# Risk Factors of Human Mpox (Monkeypox) Infection: A Systematic Review and Meta-Analysis

**DOI:** 10.1101/2024.08.14.24311975

**Authors:** Chigozie Louisa J. Ugwu, Nicola Luigi Bragazzi, Jianhong Wu, Jude Dzevela Kong, Ali Asgary, James Orbinski, Woldegebriel Assefa Woldegerima

## Abstract

**Background:** Mpox (formerly Monkeypox) virus has affected the lives of thousands of individuals both in endemic and non-endemic countries. Before the May 2022 outbreak, Mpox infections were sporadically endemic in Central and Western Africa, still research into Mpox has been limited and lacking epidemiological data. Thus, identification of potential risk factors to better understand who is at risk of being infected is critical for future prevention and control.

**Objective:** To synthesize comprehensive evidence on risk factors associated with human Mpox transmission both in endemic and non-endemic countries from inception to March 31, 2024.

**Methods:** The Preferred Reporting Items for Systematic Reviews and Meta-Analyses (PRISMA) guidelines were followed in conducting the systematic review. Electronic databases were searched. Two reviewers sifted the articles that were included in the review: firstly, by title and abstract, and secondly, by full text. We used the Newcastle-Ottawa Scale (NOS) to assess the risk of bias for included articles. Fixed or random effects meta-analysis were conducted when at least two studies reported odds ratios (OR), relative risks (RR), with 95% confidence intervals (CI). Heterogeneity was assessed using the *I*^2^ statistic and sensitivity analysis was also done. The study protocol has been registered under PROSPERO with ID: **CRD42023459895**.

**Results:** 947 articles were identified from the database search and 31 articles were eligible to be included in the systematic review. The findings of the meta-analysis showed that interaction with infected animals (*OR* = 5.61, 95% *CI* = 2.83, 11.13), HIV (*OR* = 4.46, 95% *CI* = 3.27, 6.08), other STIs (*OR* = 1.76, 95% *CI* = 1.42, 2.91), sexual contact/activities (OR = 1.53, 95% CI = 1.13, 4.82), contact with an infected person (OR = 2.39, 95%CI = 1.87, 3.05), being identified as men who have sex with men (MSM) (OR = 2.18, 95%CI = 1.88, 2.51), and having multiple sexual partners Mpox (OR = 1.61, 95%CI = 1.24, 2.09), were associated with an increased risk of contracting Mpox. However, patients who were vaccinated against smallpox had a lower risk of Mpox infection (OR = 0.24, 95%CI = 0.11, 0.55).

**Conclusion:** This study is the first meta-analysis on reported risk factors for Mpox. Our analysis demonstrated that certain factors were associated with increased risk of Mpox, whereas smallpox vaccination had a protective role against contracting Mpox. The study findings could facilitate future strategic public health planning and targeted intervention.

**Key messages of this article:** *What is already known on this topic:*

- Mpox (monkeypox) is a zoonotic infectious disease of notable global public health importance due to recent outbreaks in non-endemic countries.
- Prior outbreaks of Mpox have been associated with travel to endemic areas in Western and Central Africa, contact with infected animals, and close contact with infectious lesions, particularly among household members.

*What this study adds:*

- This study is the first meta-analysis on reported risk factors for Mpox. Our study findings add to the body of evidence on Mpox research efforts and could assist in future Mpox global strategic intervention and control.
- Our meta-analysis revealed a strong correlation between increased risk of Mpox infection, HVI, other STIs, physical and sexual contacts, and being identified as MSM.
- While HIV infection may be a risk factor for Mpox, Mpox lesions could also facilitate the transmission of HIV and other STIs.

*How this study might affect research, practice or policy:*

- The results of this systematic review and meta-analysis provide evidence to support policymakers in future Mpox intervention and prevention in both endemic and non-endemic countries based on identified risk factors.

## Introduction

The recent re-emergence of human monkeypox (Mpox) infection is a significant concern for global health (1). Mpox is an *Orthopoxvirus*, endemic in West and Central Africa, with a clinical presentation resembling that of smallpox (1, 2). The virus was initially identified in captive monkeys in Denmark in 1959 and later in a 9-year-old male child from the Democratic Republic of the Congo (DRC) in 1970 (2-4).

Over the past forty years, the DRC has persistently reported human Mpox cases, with the number of annual reports surpassing 1000 cases in the past twenty years (3). Since the initial human identification of Mpox in the DRC, there have been intermittent reports of outbreaks across Africa, characterized by a relatively low mortality rate.

Between 1970 and 2003, isolated cases of Mpox were reported in various African countries, including Nigeria, Côte d’Ivoire, Liberia, and Sierra Leone, among others (3, 5, 6). From 2003, human Mpox cases were reported outside Africa due to migration. For example in 2018, human Mpox cases were reported in Israel and the United Kingdom, in 2019 it was reported in Singapore, and in 2003 and 2021 Mpox cases were reported in the United States of America (3, 6, 7). These incidents underscored the potential for Mpox to spread globally through travelers from areas where the disease is endemic to non-endemic regions (3).

In May 2022, the world experienced the first human Mpox outbreaks which were largely reported in regions with no previous history of the disease and began to spread across multiple countries (1). This led the World Health Organization (WHO) to declare it a Public Health Emergency of International Concern (PHEIC) in July 2022 (1, 8). The latest data indicates over 90,000 confirmed cases of Mpox infections globally, with around 150 fatalities across 110 countries. Non-endemic countries greatly impacted were the USA, Canada, Spain, France, Colombia, the UK, Mexico, Brazil, Peru, and Germany as they represent approximately 86% of the reported global cases (1). Mpox infection has greatly declined globally as a result of several factors including public health intervention, behavioral adjustment, and an increasing level of immunity within the population at risk, either naturally acquired or via effective Mpox vaccination campaigns (9). However, Mpox still remains a major global public health concern needing critical research for future control and interventions (1, 10).

A significant aspect of the Mpox cases in 2022 was the lack of direct epidemiological ties to the importation of animals or migration, with the cases being mostly linked to sexual contacts (1). The individuals affected predominantly identified themselves as gay, bisexual, or other men who have sex with men (1, 11, 12). In contrast, in certain countries, individuals with confirmed Mpox cases reported having traveled to nations within Europe and North America (1, 13). This marks a departure from traditional transmission routes previously observed and points to a new pattern of transmission within specific communities (1). Therefore, this systematic review aims to explore all risk factors associated with the transmission of Mpox both in endemic and non-endemic countries.

## Materials and Methods

### Study Protocol

This systematic review and meta-analysis were conducted and reported following the Preferred Reporting Items for Systematic Reviews and Meta-Analyses (PRISMA) guidelines (14, 15). Additionally, the review was registered with the International Prospective Register of Systematic Reviews (PROSPERO) under the registration number CRD42023459895.

### Search Strategy

Electronic databases including Scopus, Google Scholar, Cochrane Library, Web of Sciences (WOS), EMBASE, and PubMed were searched systematically for potential publications from inception to March 31, 2024. The key search terms were: “Mpox” OR “Monkeypox” OR “Orthopoxvirus” OR “Monkeypox virus” AND (“risk factors” OR “influential factors” OR “impact factors” OR “exposure”. The search was specifically tailored to select original articles published in the English language, including any study design (case-control, observational (prospective/retrospective), and randomized control studies) with the human population. To streamline the screening process and enhance efficiency, EndNote X9 (16) was employed to keep a record of all articles obtained from the electronic database, duplicates removed, and final studies included in the review.

### Eligible Criteria

Articles included were original articles of any study design (randomized control trial, case study, observational study, etc.) that investigated the risk factors of Mpox infection in human subjects. The study must have a minimum sample size of 10 patients and be published in the English language. Articles excluded were review papers, grey literature (like dissertations and thesis), studies with less than 10 participants, and studies on animal and non-human populations.

### Study Selection

The study selection was carried out by two reviewers (CLJU and WAW) independently., The reviewers screened all articles obtained from the database (after removing the duplicates) initially by title and abstract, then by full text. The eligible criteria were used in the screening process. Disagreements were resolved through discussion and mutual agreement and consensus were reached.

## Statistical Analysis (Meta-Analysis)

Meta-analyses were conducted for risk factors that were reported in two or more studies and included the estimated odds ratios (ORs), or relative risks (RRs) (17). The degree of heterogeneity was determined quantitatively using *I*^2^ index statistic (18, 19). *I*^2^ ≥ 75% is a measure of highly significant heterogeneity, *I*^2^ = 50% − 70% is recognized as moderate heterogeneity, 25% < *I*^2^ < 50% denotes a low heterogeneity and *I*^2^ ≤ 25% presents homogeneity. Thus, heterogeneity is high when Cochran’s Q *p* − *value* < 0.10, and *I*^2^ ≥ 50% (20, 21). For a better-quality study, we employed the fixed-effects meta-analysis model when heterogeneity was low or homogeneous, opted for the random-effects model if it was highly significant, and conducted sensitivity analysis to identify the sources of heterogeneity (17). To explore possible publication bias, funnel plots with Egger’s weighted regression test were used (20). All of the analyses were implemented in the R statistical software version 4.3.2 with R-package *meta* (22, 23). *p* − *value* < 0.05 were considered statistically significant.

### Data Extraction

Authors extracted the following data from the included studies: the author’s name, country of study, year of publication, study design, sample size, demographic variables (such as gender, age, sexual orientation, etc.), clinical variables (such as disease presentations, etc.), outcome (Mpox), exposure (risk factors), and odds ratios, or relative risks plus the 95% CIs in an Excel spreadsheet.

### Quality Assessment

The evaluation of the study’s quality was conducted using the Newcastle-Ottawa Quality Assessment Scale (NOS) (24, 25). The NOS framework examines several aspects: (1) Selection, focusing on the adequacy of case definition, representativeness of cases, selection of controls, and definition of controls. (2) Comparability, assessing whether cases and controls are comparable based on the study’s design or analysis. (3) Exposure, checking the ascertainment of exposure, consistency in the method of ascertainment for cases and controls, and the non-response rate. In this study, articles that achieved Newcastle-Ottawa Scale (NOS) scores ≥ 5 were considered high-quality publications.

## Results

### Literature Search and Study Selection

Initial searches identified 947 studies, and 8 additional studies were identified from citation referencing. Subsequently, 460 studies were obtained after removing duplicate literature, and 291 were excluded after screening titles and abstracts. After a full-text review, a total of 138 texts were further excluded for not meeting the inclusion criteria. Finally, 31 studies were included in the systematic review (**Figure 1)**.

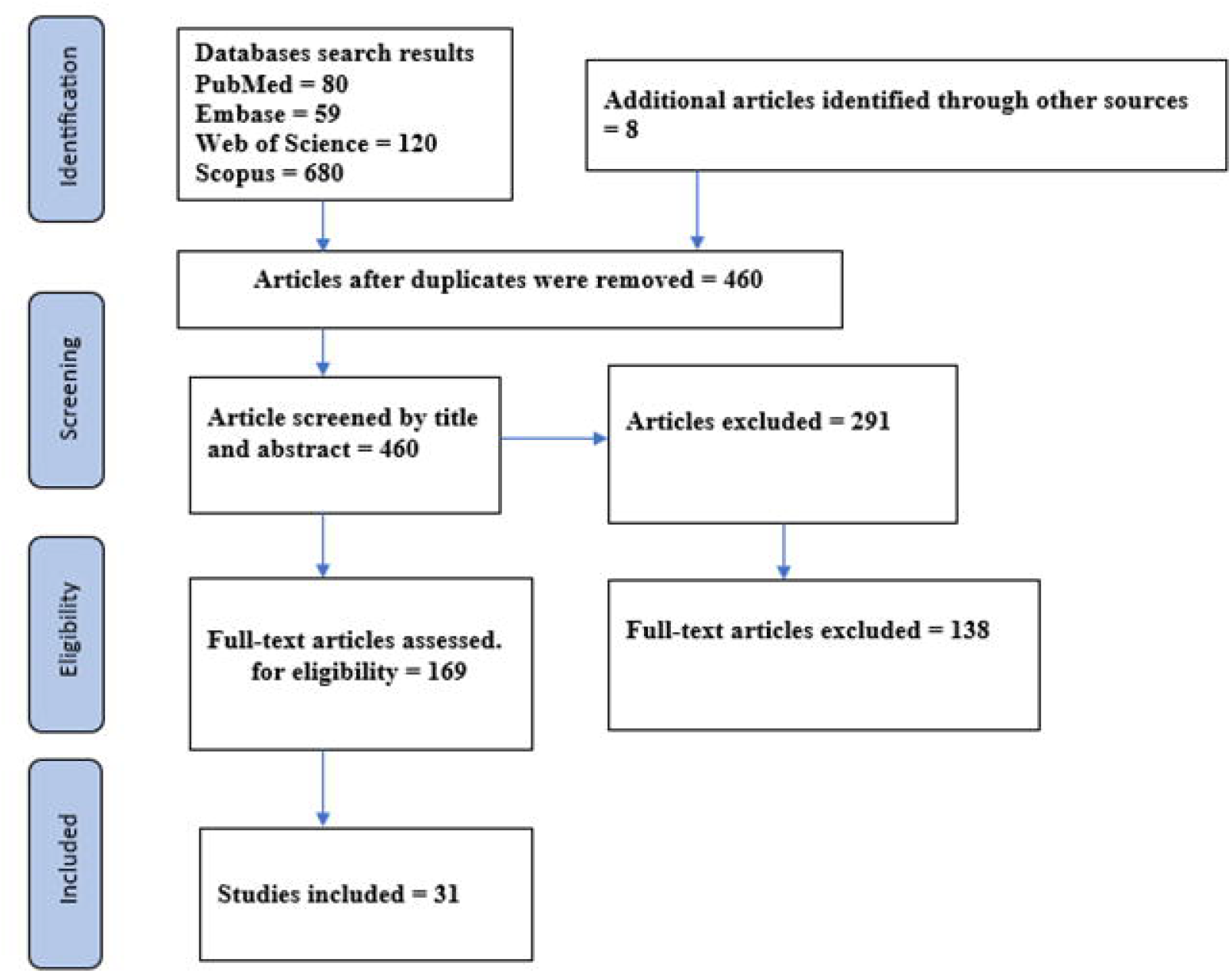

### Demographic characteristics of the included studies

Of the 31 included studies in this systematic review, seven studies were conducted in DR Congo (26-31), one in Nigeria (6), two in the USA (32, 33), two in the UK (34, 35), four in Spain (36-39), two in Brazil (40, 41), four in Italy (42, 43), two in UAE (26, 44), one in the Netherlands (45), one in Israel (46), one in Chile (47), one in Portugal (48), one in Belgium (49), one in Germany (50), and four multi-country studies for a group of countries (34, 51, 52) (see **Figure 2)**. The data included 148,499 Mpox cases involving men, women, cisgender, transgender, and non-binary individuals. A summary of the study characteristics is reported in **Table 1**.

**Table 1:** Demographic characteristics of the study population (Mpox confirmed cases)

| No | Author (year) | Country | Study design | Sample size | Mean age | Sex (M/F) | Sexual Orientation (MSM/gay/bisexual) |
| --- | --- | --- | --- | --- | --- | --- | --- |
| 1 | Acevedo, A. (2022) (47) | Chile | Cases-control study | 1415 | 35 years | Male | - |
| 2 | Akilimali, A. (2022) (26) | DR Congo | Cases-control study | 2159 | 5+ | Male/Female | MSM |
| 3 | Alegre, B. (2022) (36) | Spain | Cohort study | 11 | 35 ± 8 years. | Male | MSM |
| 4 | Alhammadi, O. A. (2022) (53) | UAE | Prospective cohort study | 176 | 30.4 ±7 years | Male (164) | - |
| 5 | Alpalhão, M. (2023) (48) | Portugal | Prospective cohort study | 42 | 37 years | Male | MSM |
| 6 | Angelo, K. (2022) (34) | 15 countries | Cohort study | 226 | 37 (18–68; 32–43) | Male (211) | MSM, Gay, Bisexual (208 of 211) |
| 7 | Berens-Riha, N. (2022) (49) | Belgium | Cohort study | 169 | 39 years | Male/Female | - |
| 8 | Candela, C (2022) (42) | Italy | Observational study | 140 | 37 years | Male (137) | MSM (134 of 137) |
| 9 | Catala A. (2022) (37) | Spain | Prospective cohort study | 185 | 37 years | Male | MSM: All reported having sex with men. Having multiple sexual partners during the previous weeks. |
| 10 | Ciccarese, G. (2022)(43) | Italy | Cases-control study | 16 | 37 years | Male | Homosexual and bisexual |
| 11 | Cline, A. (2022) (32) | USA | Cross-sectional study | 250 | 30-60 years | Male (242)<br>Female (7) | - |
| 12 | Dar, N. G. (2023) (44) | UAE | Cases-control study | 16 | 33.9year | Male | - |
| 13 | Doshi, R. H. (2017) (27) | DR Congo | Observational study | 43 | 11.5 years | Male<br>Female (11) | - |
| 14 | Estévez S.(2022) (38) | Spain | Observational study | 133 | 33 | Male | - |
| 15 | García-Piqueras (2022) (39) | Spain | Cohort study | 53 | 36 years | Male | MSM (49) |
| 16 | Laurenson-Schafer, H (2023) (54) | WHO States | Observational study | 76, 293 | 30-45 years | Male | MSM (29,854) |
| 17 | Lin, M. (2023) (51) | 39 nations | Observational study | 30, 012 | 30 years | Male and Female | - |
| 18 | Martins-Filho, P. R (2022) (40) | Brazil | Observational study | 9729 Deaths (12) | 20–39 years | Male | MSM |
| 19 | Nolen, L. D (2015) (28) | DR Congo | case-control study | 13 | 35 years | Male and Female | - |
| 20 | Oeser, P. (2022) (55) | Germany | Cross Sectional Study | 1840 | 18-59 years | Trans and non-binary | MSM (trans and non-binary) |
| 21 | Ogoina, D. (2023) (6) | Nigeria | Cohort study | 160 | 35 years | Male (114) and Female (46) | - |
| 22 | Quiner, CA. (2016) (29) | DR Congo | Observational study | 939 | 8+ years | Male (608) and female (331) | - |
| 23 | Reynolds, Mary G (2003)(33) | USA | Case-control study | 61 | 18+ years | Male and female | - |
| 24 | Rimoin, AW. (2007) (30) | DR Congo | Observational study | 760 | 11.9 years | Male and Female | - |
| 25 | Souza I. (2023) (41) | Brazil | Observational study | 10,169 | 15+ years | Male | MSM |
| 26 | Thornhill, J. (2022) (52) | 16 countries | Case-control study | 528 | 38 years | Male | MSM |
| 27 | Vallejo-Plaza (2022) (56) | Spain | Cohort study | 158 | 16–76 years | Female | - |
| 28 | van Ewijk (2022) (45) | Netherlands | Observational study | 1000 | 31-45 years | Male | MSM |
| 29 | Vivancos, Roberto (2022) (35) | UK | Case-control study | 86 | 38 years | Male/female | MSM |
| 30 | Whitehouse, E, (2015) (31) | DR Congo | Observational study | 3639 | 14 years | Males and Females | - |
| 31 | Zucker R. (2022) (46) | Israel | Cohort study | 8088 | Aged 25 to 46 | Male | - |

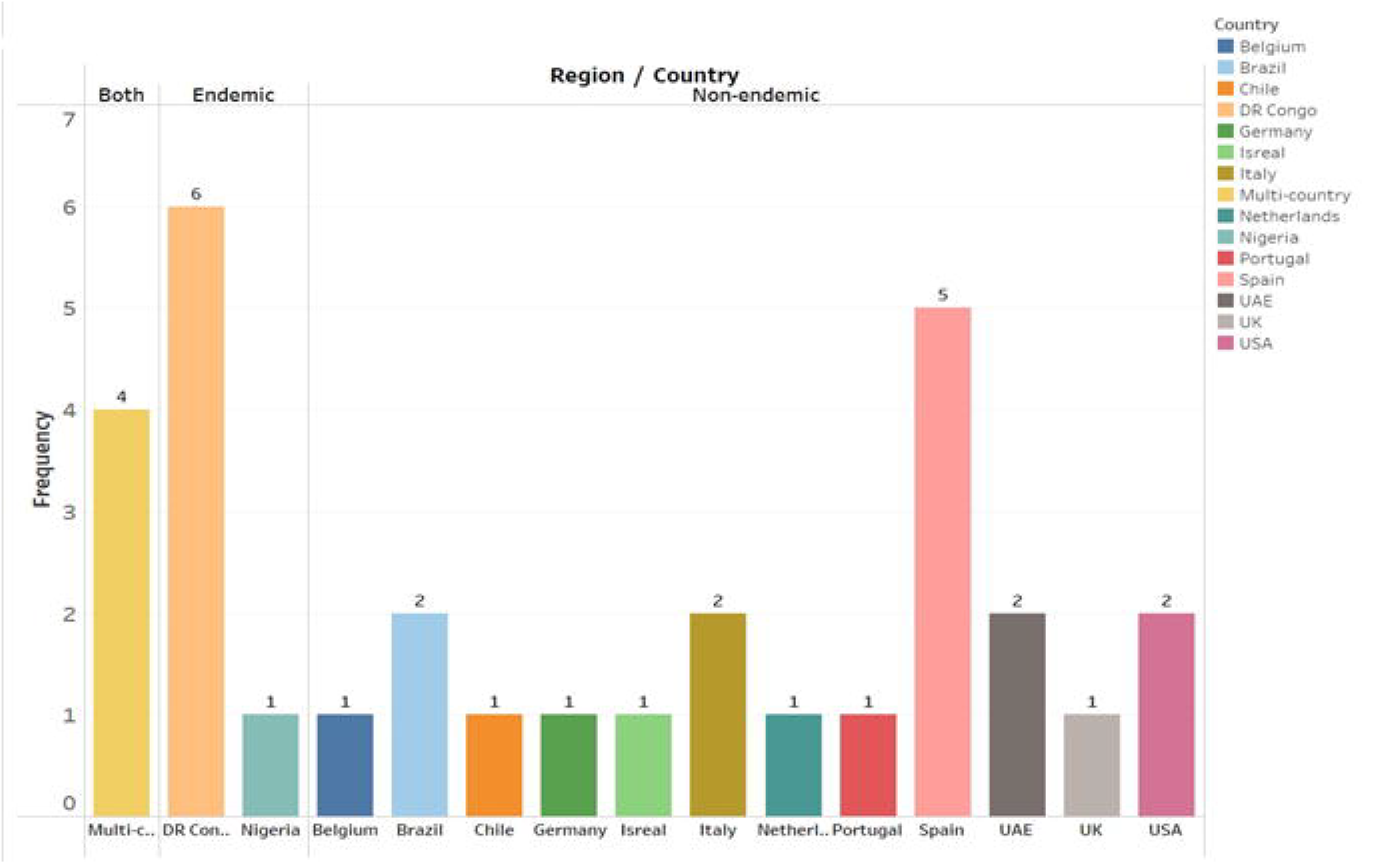

### Clinical Characteristics of the Included Studies

All risk factors and clinical presentations identified in 31 included articles are reported in **Table 2**. The most common symptoms of Mpox are fever, rash, exanthema, lymphadenopathy, myalgia, lesions, and headache, with fever being the predominant systemic manifestation observed (6, 38, 47). **Figure 3** summarizes the clinical symptoms of mpox reported in the papers included in the review. Individuals with Confirmed Mpox infection were mostly presented with mucocutaneous lesions, most commonly on the genital and anal areas, which support sexual contact as a means of Mpox transmission (6, 47, 52).

**Table 2:**
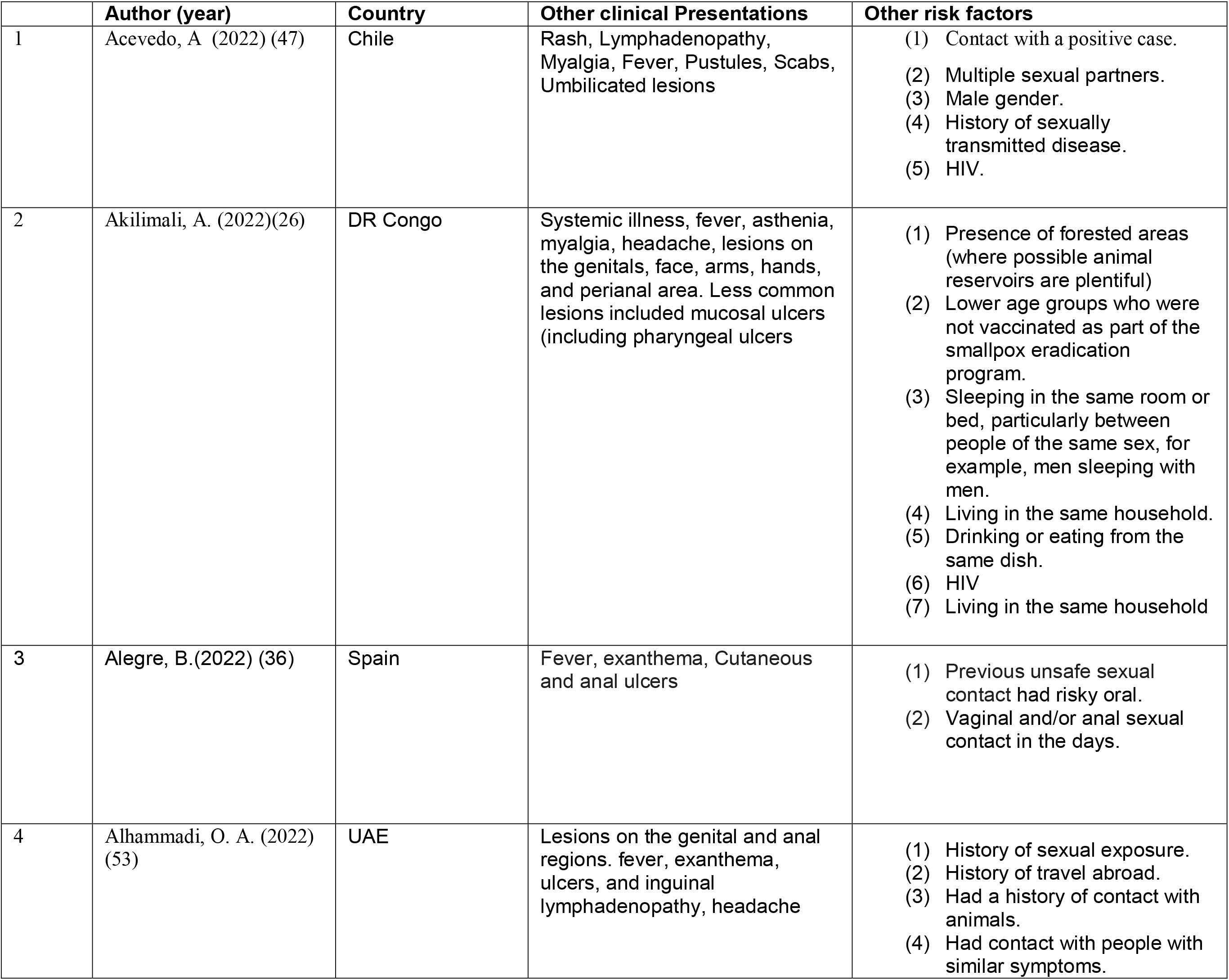
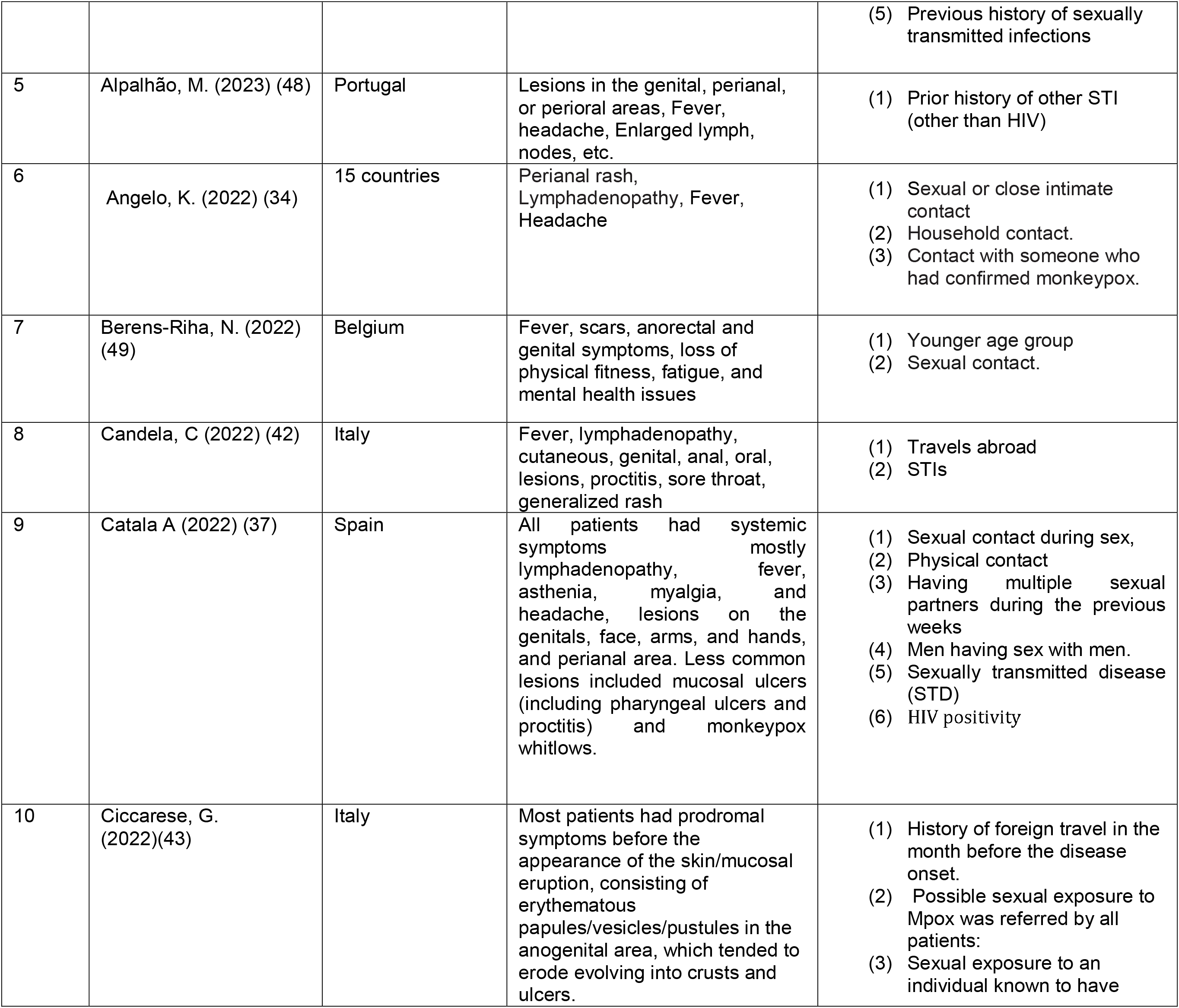
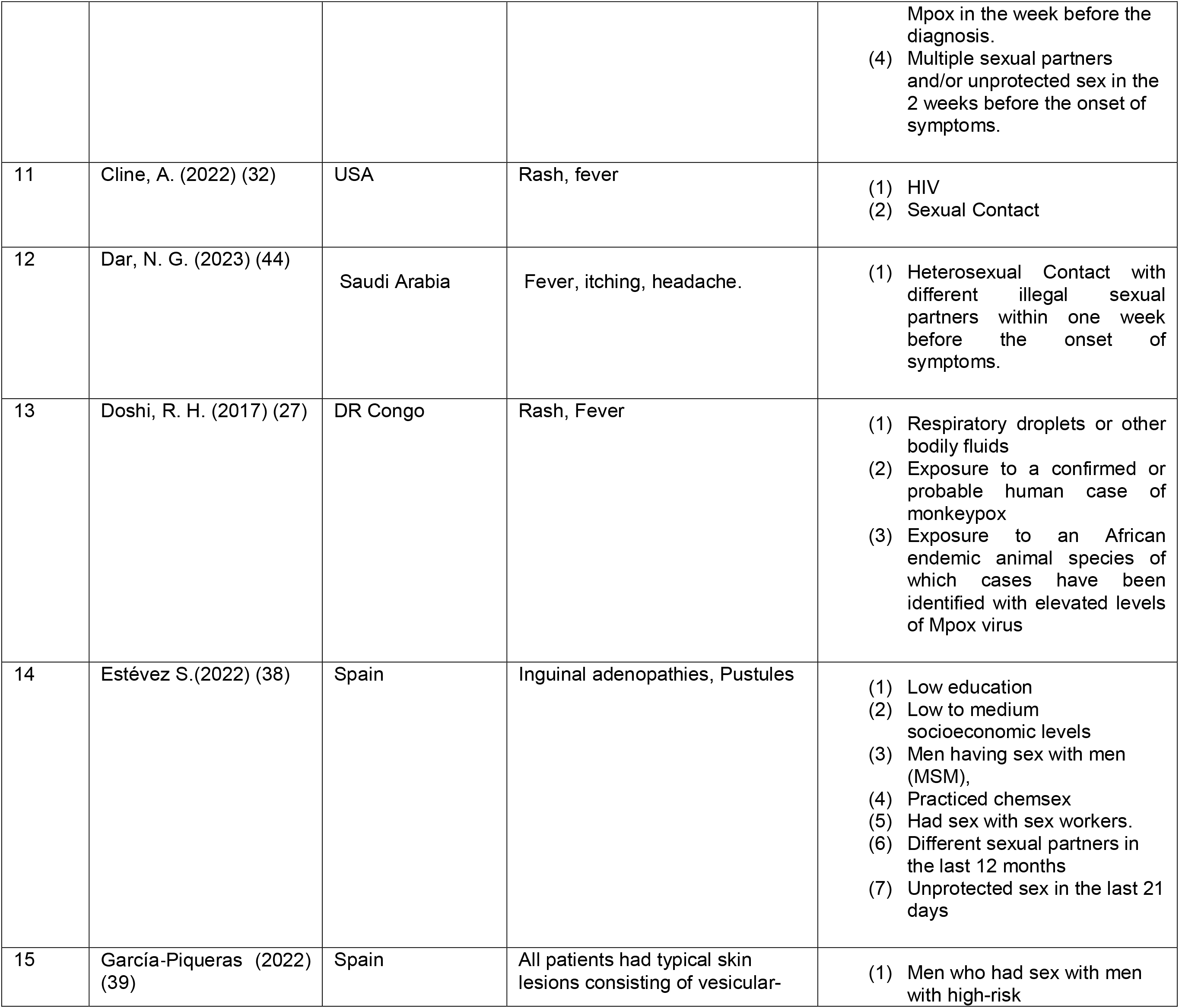
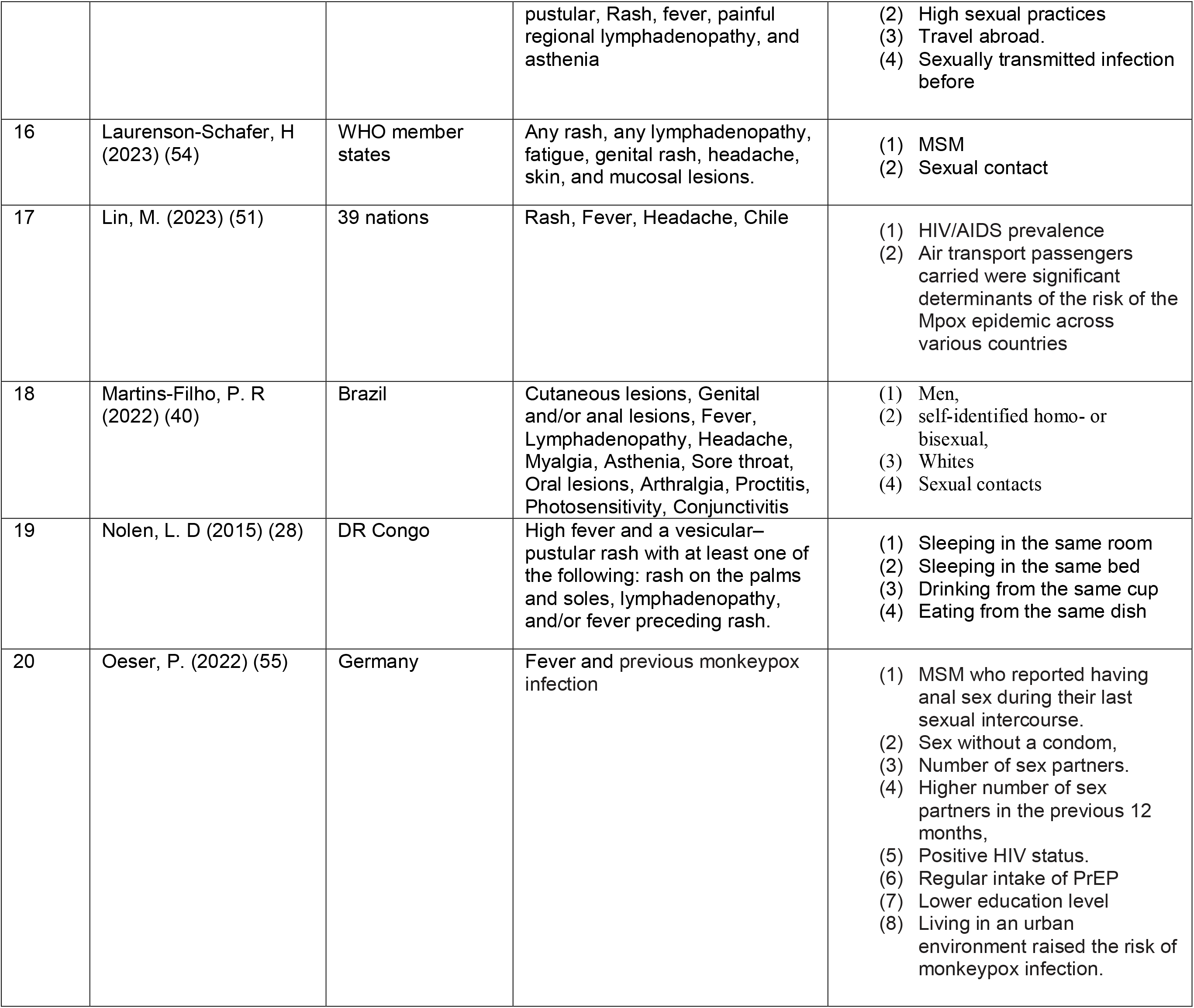
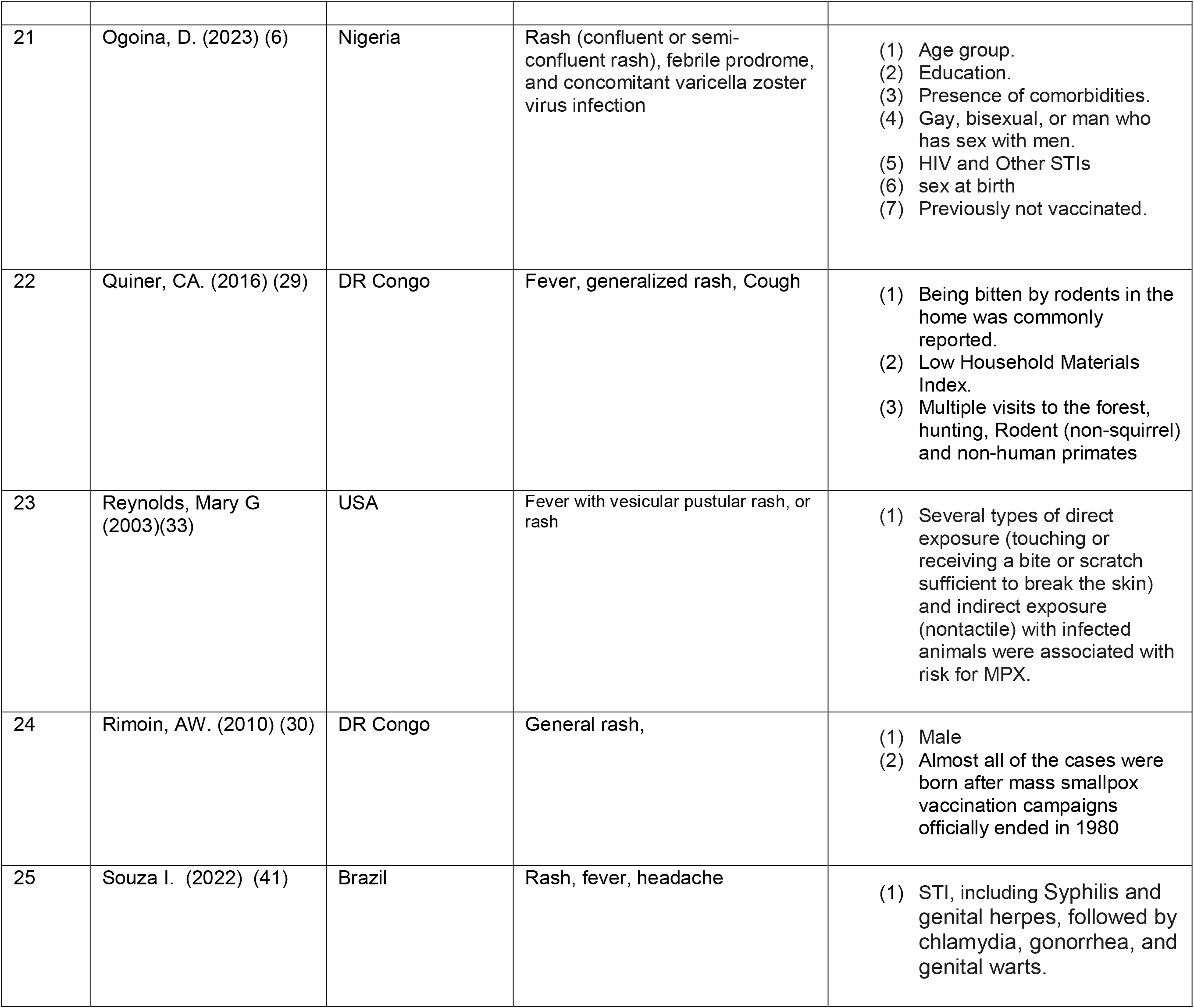
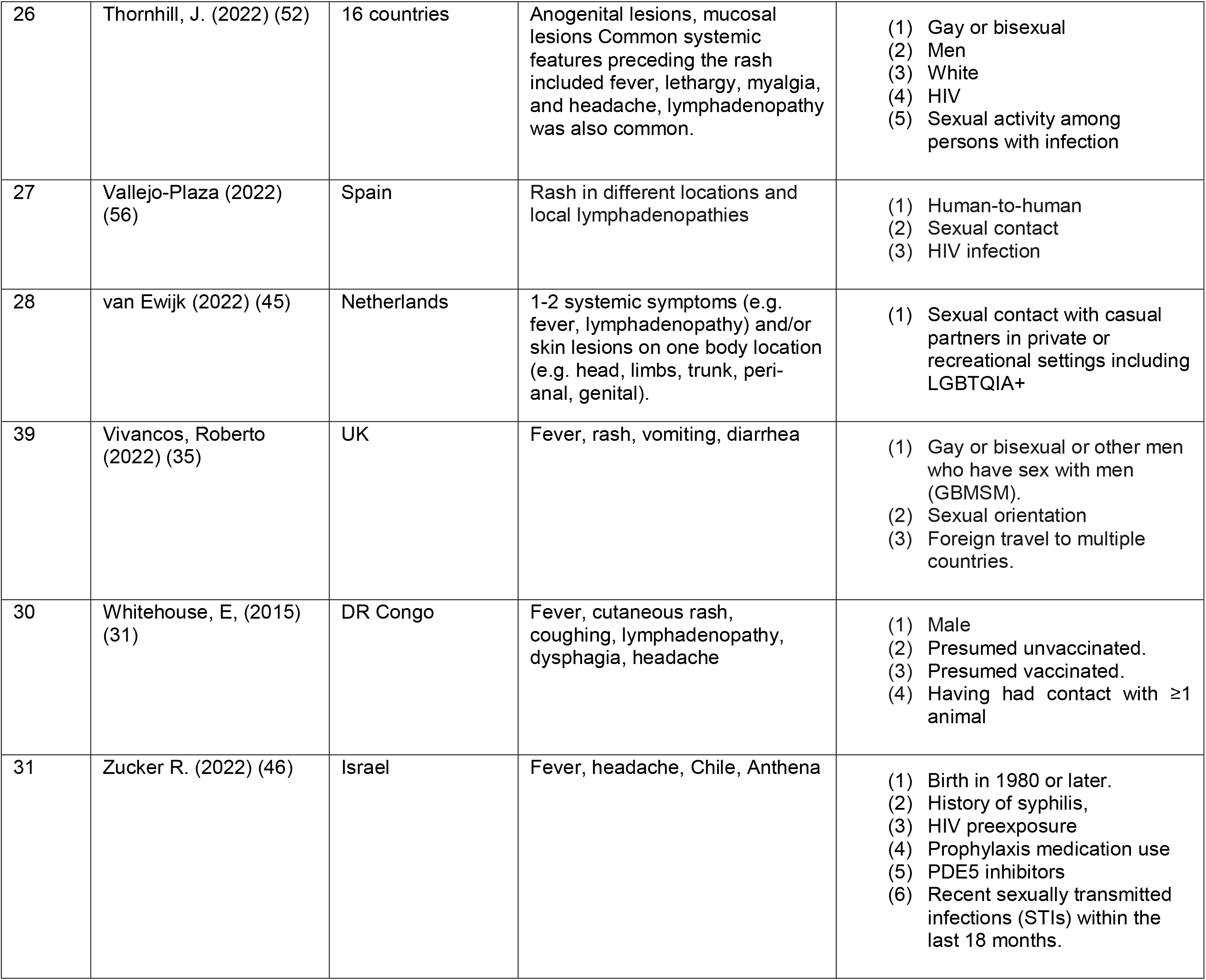
Clinical characteristics of patients, and identified risk factors of Mpox.

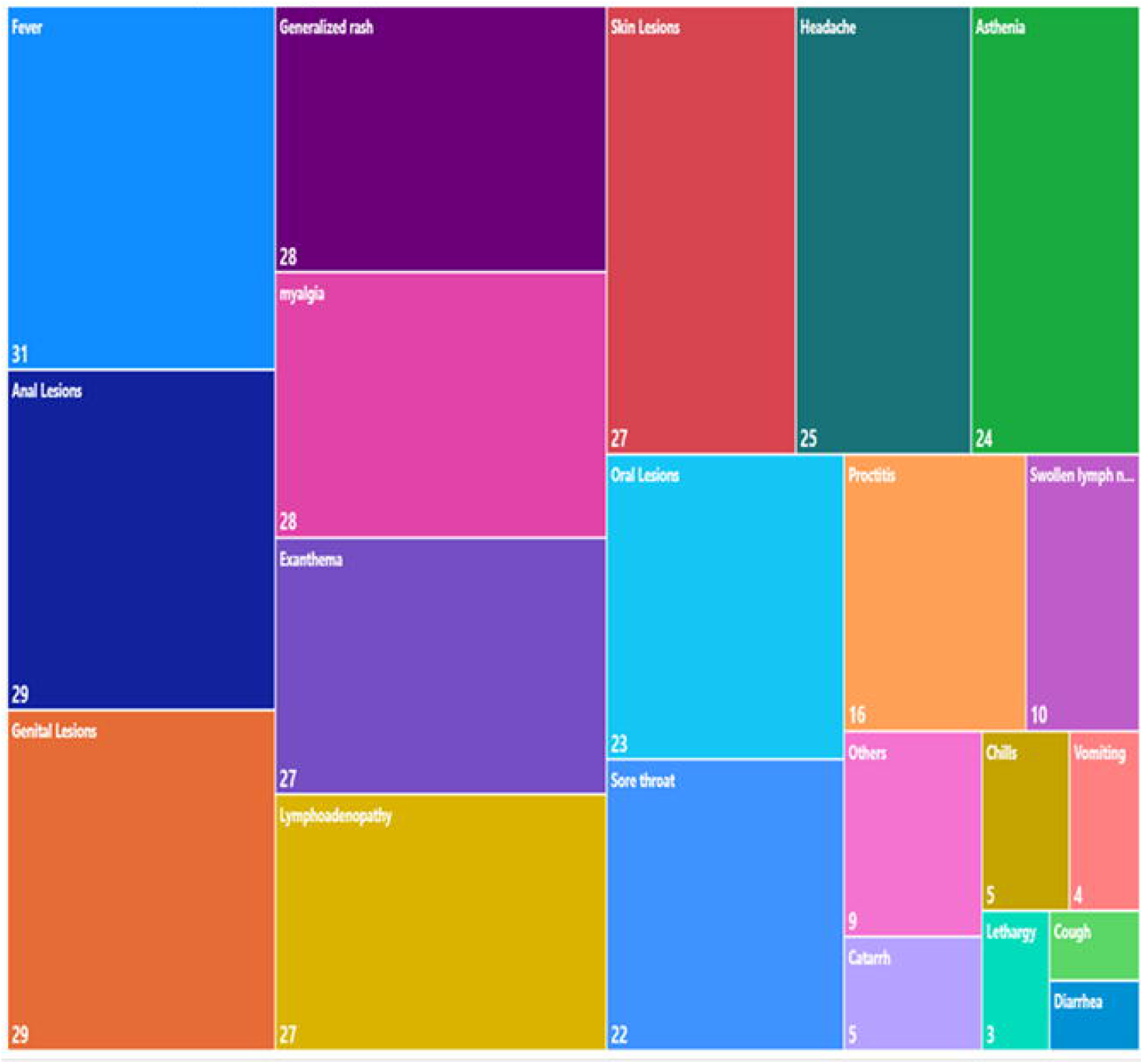

The STIs reported among Mpox-infected patients included HIV – being the most prevalent (34, 38, 47, 51, 52, 54). This may be likely due to the relative immunodeficiency associated with HIV infection, even among those receiving treatment (43). Though HIV infection highly increases the risk of contracting Mpox, Mpox lesions could also potentially enhance the transmission of HIV and other STIs (30, 47, 53). A summary of STIs reported in the included studies is presented in **Figure 4**.

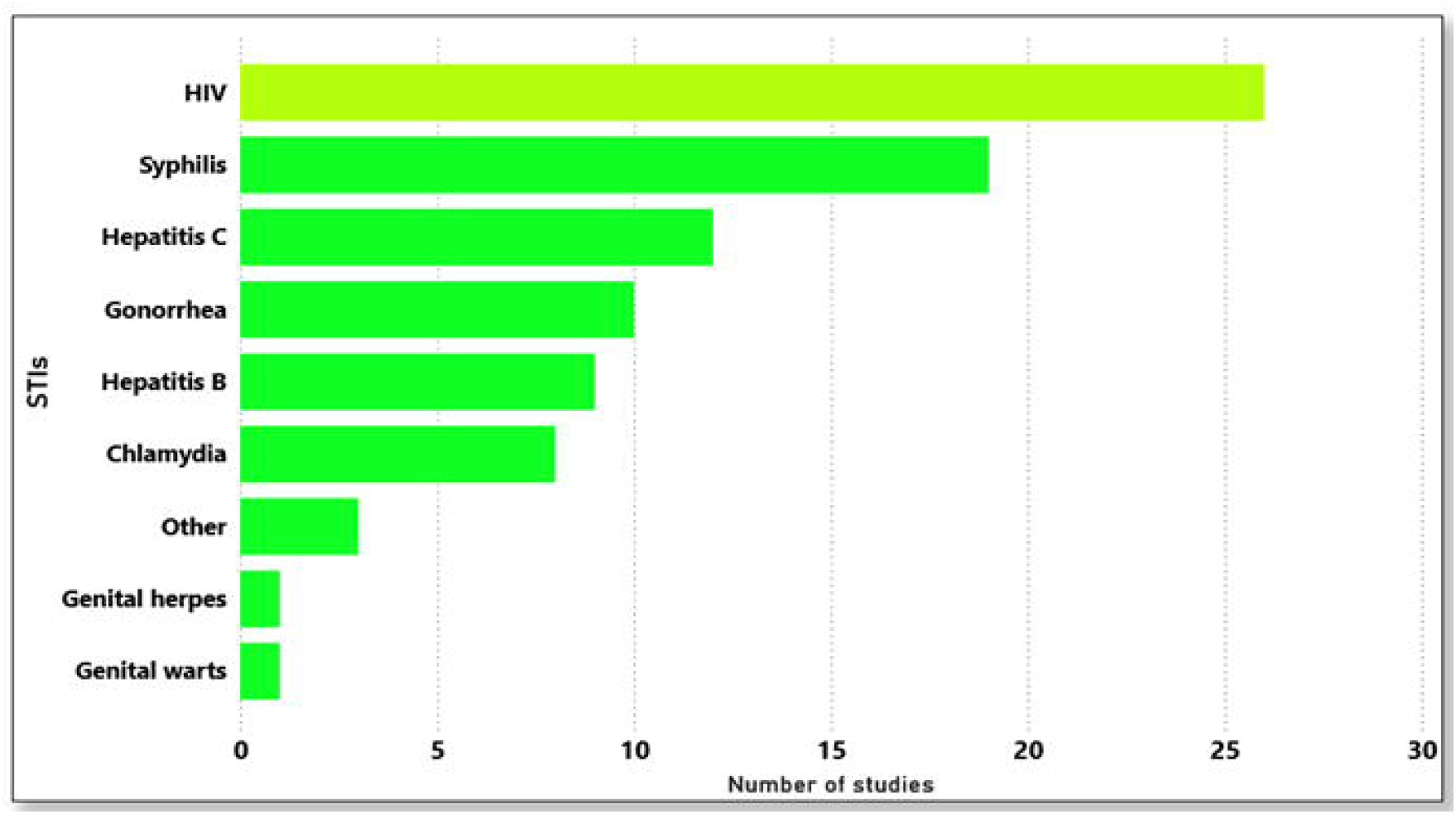

## Meta-analysis of Risk Factors of Mpox

### Interaction with Animals

Among the included studies, two studies in endemic regions (29, 33) identified close contact with wild animals, daily exposure during an animal’s illness, contact with rashes or eye crusts, scratching, cleaning cages or handling bedding, and direct exposure to animals susceptible to Mpox infection. Our study has detected a moderate heterogeneity (*I*^2^ = 42%, *p* = 0.16). Therefore, the fixed-effects model was adopted for meta-analysis. From the result of the forest plot in **Figure 5**, interaction with animals was found to be a statistically significant risk factor for Mpox in endemic areas (*OR* = 5.61, 95%*CI* = 2.83, 11.13, *P* − *value* < 0.0001). These findings reveal that being bitten by rodents at home, handling Mpox-infected animals, and daily exposure to their excretions and secretions were statistically and significantly linked to higher rates of human Mpox infection. The results indicate that individuals with direct exposure to infected animals are 5.61 times more likely to contract Mpox than those without such exposure (**Figure 5**).

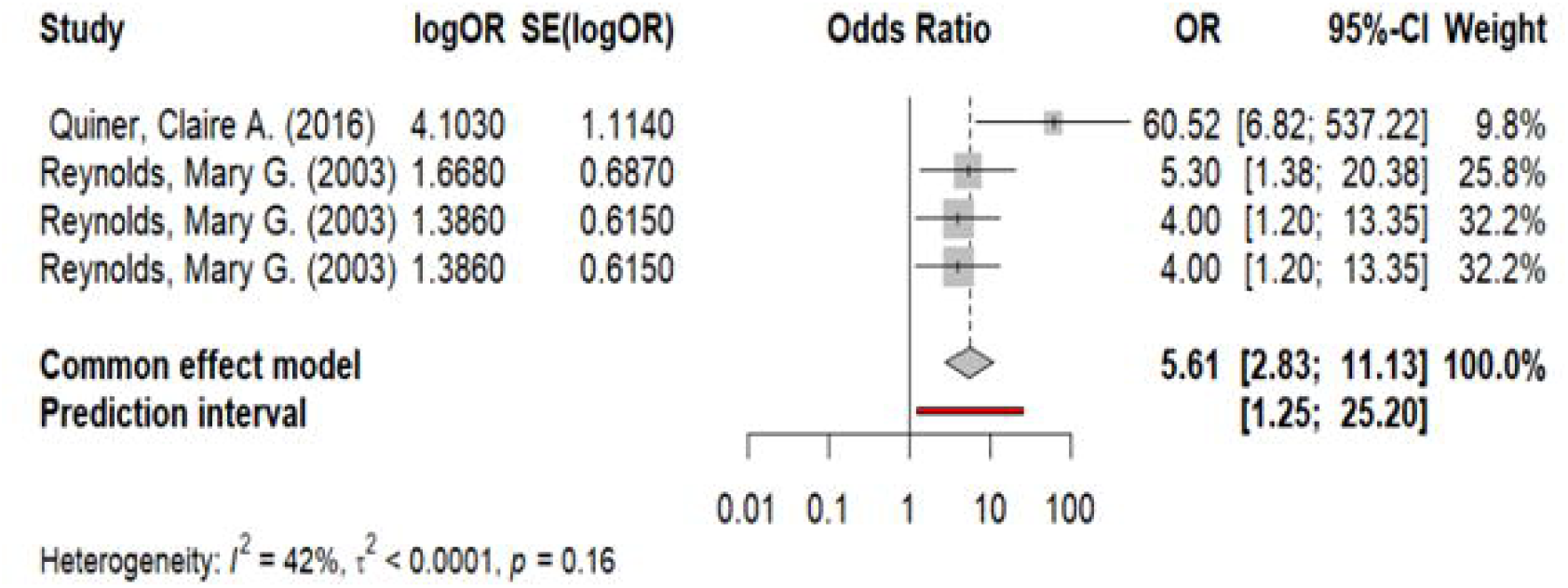

### Human Immunodeficiency Virus (HIV)

Five studies 6, 46, 47, 51, 55), identified HIV as both a risk factor and a comorbidity for Mpox infection. Individuals with advanced HIV (who are immunocompromised) have an increased risk of severe Mpox symptoms and mortality. Significant high heterogeneity was detected (*I*^2^ = 87%, *p* < 0.01), and a random-effects model was employed for meta-analysis of the association between HIV and Mpox infection. The estimate obtained showed that HIV is a risk factor for Mpox (*OR* = 4.05, 95%*CI* = 2.02, 8.14, *P* − *value* < 0.0001). Individuals living with HIV are 4 times more likely to contract Mpox compared to those without HIV, underscoring the role of HIV as a significant risk factor for Mpox infection (**Figure 6**).

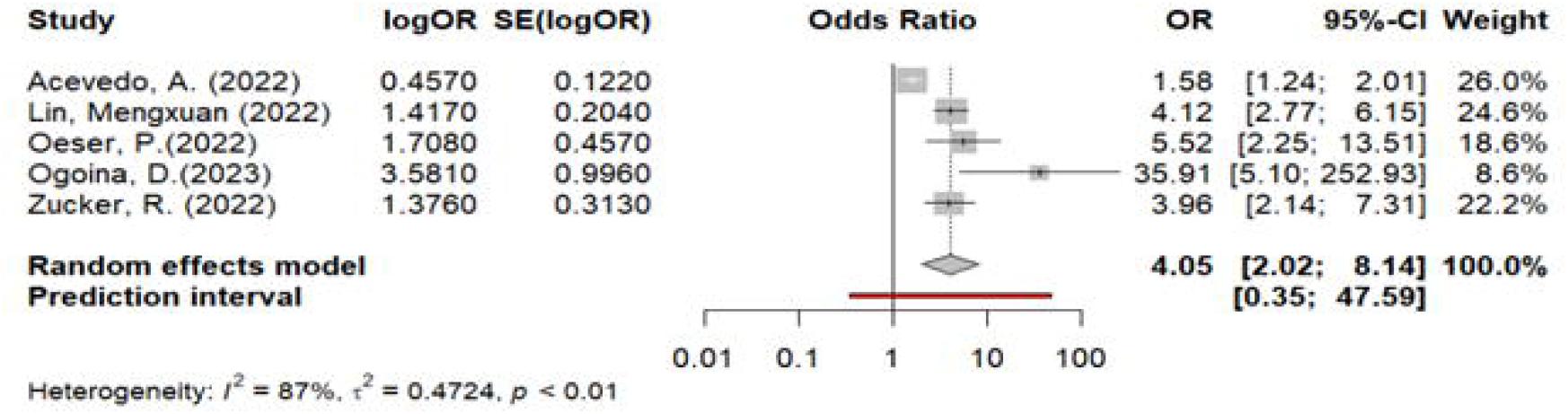

However, to identify the source of high heterogeneity, we conducted a sensitivity analysis and detected that Acevedo, A. et al. (47) was the source of heterogeneity. Removing the study reduced the heterogeneity to (I^2^ = 39%, p = 0.18). Subsequently, a fixed effect model was performed for meta-analysis. Our findings still indicate that HIV is a statistically significant risk factor for Mpox (OR = 4.46, 95%CI = 3.27, 6.08, *P* - *value* < 0.0001), suggesting that individuals with HIV are 4.5 times more likely to contract Mpox compared to those without HIV (**Figure 7**).

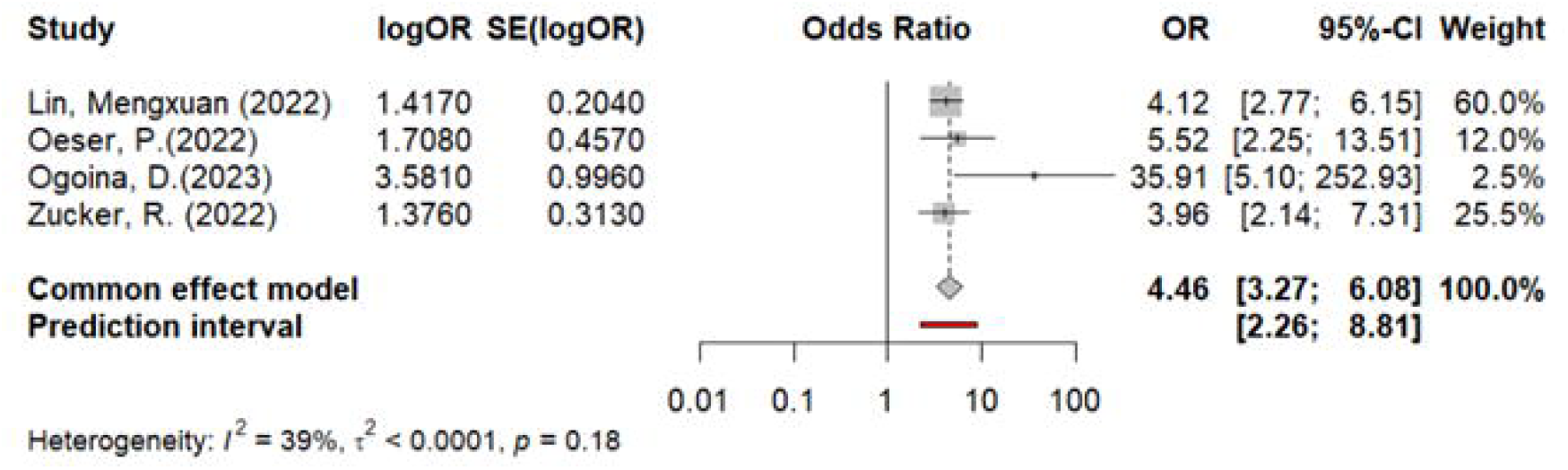

### Other Sexually Transmitted Diseases (STIs)

Three studies (6, 46, 47), reported syphilis, concomitant varicella-zoster virus infection, chlamydia, hepatitis B and C, gonorrhea, and histories of other STIs as risk factors for Mpox. From the forest plot, an evident moderate heterogeneity was detected (*I*^2^ = 40.3%, 0.19), and a fixed-effects model was performed for meta-analysis. The results indicated that other STIs are risk factors for Mpox (*OR* = 1.76, 95%*C1* = 1.42, 2.91, *P* − *value* < 0.0001), **Figure 8**.

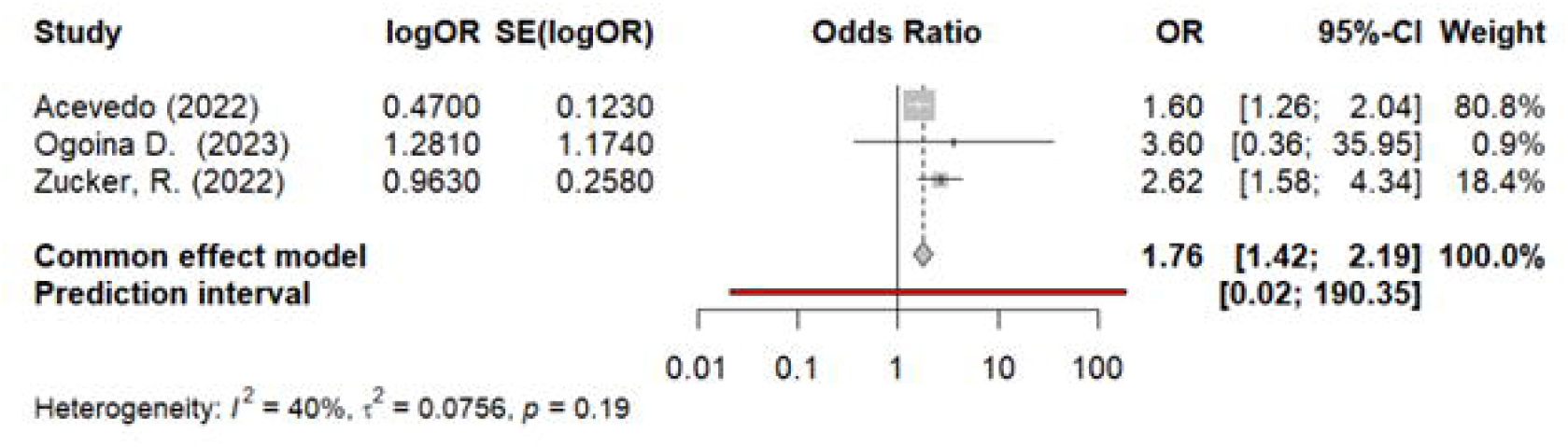

### Presence of Comorbidities

Three studies (6, 45, 47), identified the presence of comorbidities as a risk factor for Mpox infection. The forest plot analysis revealed no evidence of heterogeneity (*I*^2^ = 0%, 0.78), and a fixed-effects model was utilized for the meta-analysis. The result indicated that commodities were a risk factor for Mpox (*OR* = 1.58, 95%*CI* = 1.31, 1.91, *P* − *value* < 0.0001), **Figure 9**. However, the studies did not specify the types of comorbidities involved, leaving it unclear which specific comorbidities have a greater impact on Mpox infection.

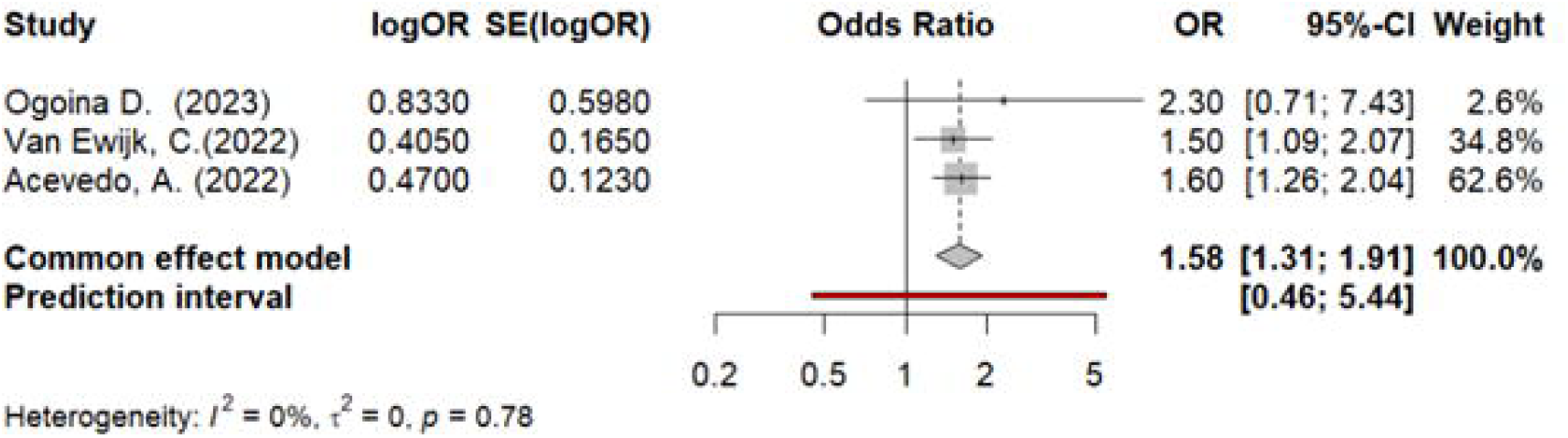

### Men Who Have Sex with Men (MSM)

Two studies (41, 45), identified specific risk factors for Mpox infection among a broader population defined as MSM, which includes transgender and non-binary individuals. These studies indicated that the majority of Mpox cases predominantly affected MSM men, with a high incidence of lesions occurring in the anogenital area. With no heterogeneity detected (*I*^2^ = 0%, 0.39), and a fixed-effects model was selected for meta-analysis. Our results indicate that MSM individuals are at a higher risk of contracting Mpox (OR = 2.18, 95%CI = 1.88, 2.51, P − value < 0.0001), **Figure 10**, with 2.18 times greater odds compared to those who are not MSM.

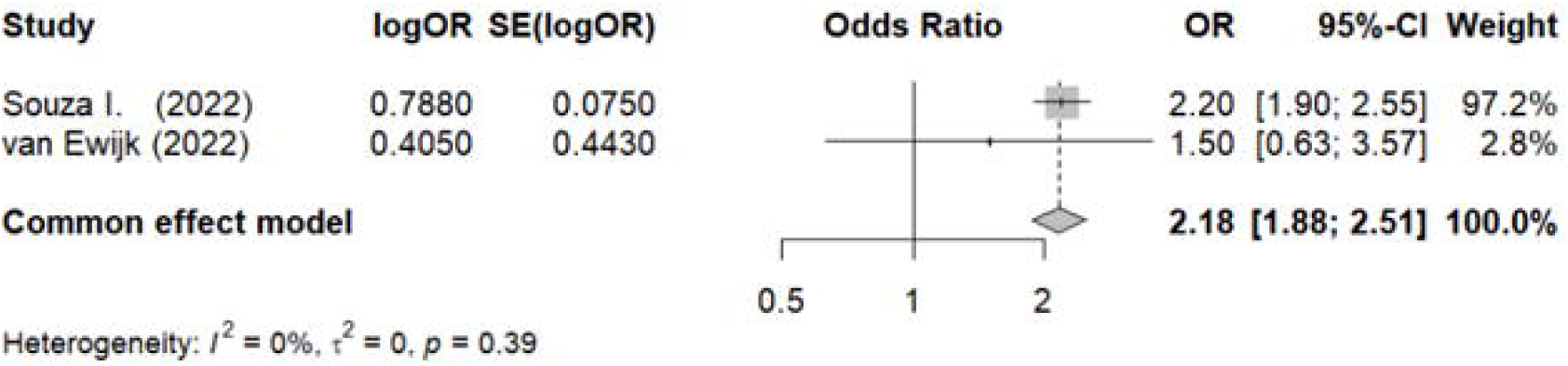

### Sexual Contact/Activities

Two studies (45, 55) reported that unprotected anal sex, or engaging in three or more sexual activities, such as oral, anal, and/or oral-anal, compared to just one or two activities in the 21 days before symptom onset, increased the risk of Mpox infection. No heterogeneity was detected (*I*^2^ = 0%, 0.74), and a fixed-effects model was selected for meta-analysis. From the forest plot, sexual contact was a risk factor for Mpox (OR = 1.53, 95% CI = 1.13, 4.82, P = 0.005). The result indicates that those who engaged in unprotected anal and oral sex had 1.53 times the risk of Mpox compared to those who did not (**Figure 11**).

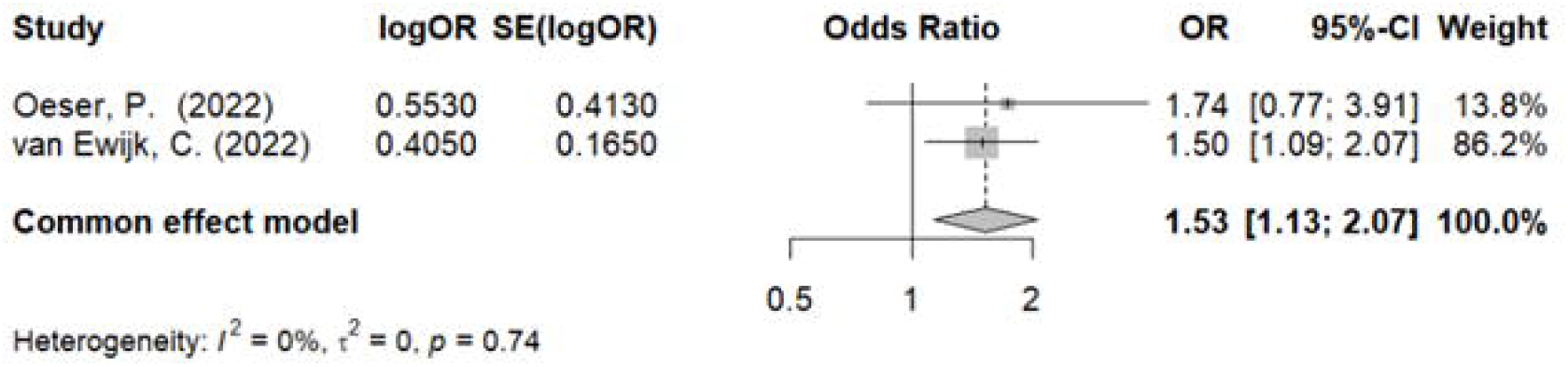

### Multiple Sexual Partners

Four studies (41, 45, 47, 55) reported having multiple sexual partners as a risk factor for Mpox infection. We analyzed the data and found significant heterogeneity (*I*^2^ = 92%, *p* < 0.01), and a random-effects model was performed for the meta-analysis. The results indicated that having multiple sexual partners is a risk factor for Mpox (OR = 1.79, 95%CI = 1.04, 3.11, P − value < 0.0001), **Figure 12**.

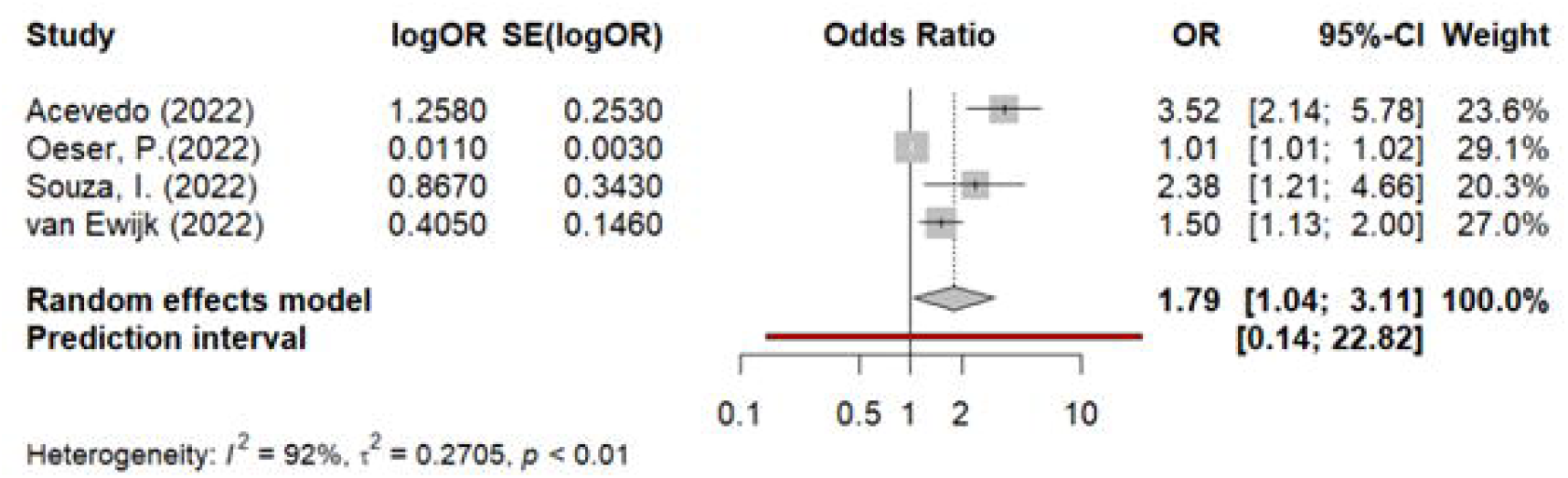

To find the source of heterogeneity, we performed sensitivity analysis and found that two studies (47, 55), were the source, and the heterogeneity was significantly reduced to (*I*^2^ = 35%, 0.22), after removing the studies. Finally, a fixed-effects model was employed for the meta-analysis, and having multiple sexual partners was concluded as a risk factor for Mpox (OR= 1.61, 95%CI = 1.24, 2.09, P < 0.0001), **Figure 13**. The result indicated that those who engage in multiple sex have about 1.6 times higher risk of contracting Mpox compared to those who do not.

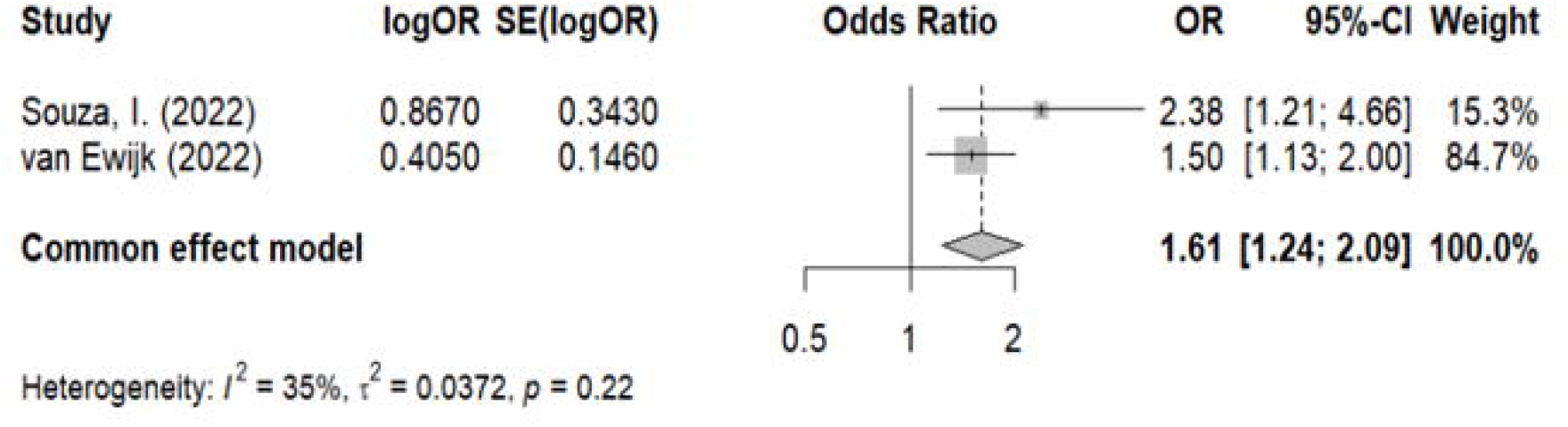

### Contact with an Infected Person

Three studies (45, 47, 49), reported previous close contact with a confirmed case, including sharing personal items like glasses and towels as a significant risk factor for Mpox. High heterogeneity was detected (I^2^ = 79%, p < 0.01), and a random-effects model was employed for meta-analysis. The result confirmed that close contact with an infected person is a risk factor for Mpox (OR = 2.05, 95%CI = 1.10, 3.79, P − value < 0.0001), **Figure 14**.

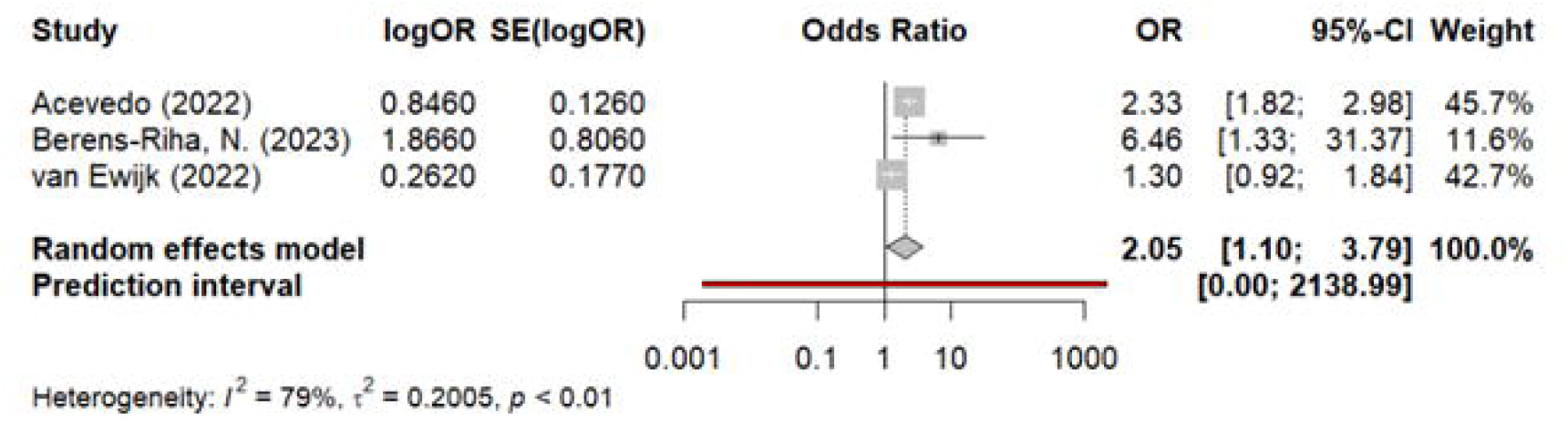

Based on the results of the sensitivity analysis, the study by van Ewijk, Catharina E. et al. (45), was identified as the source of heterogeneity. Upon its removal, heterogeneity decreased (*I*^2^ = 36%, 0.21), allowing for the use of a fixed-effects model in the meta-analysis. The analysis concluded that close contact with infected individuals significantly increases the risk of Mpox, with (OR = 2.39, 95%CI = 1.87, 3.05, P − value < 0.0001), **Figure 15**.

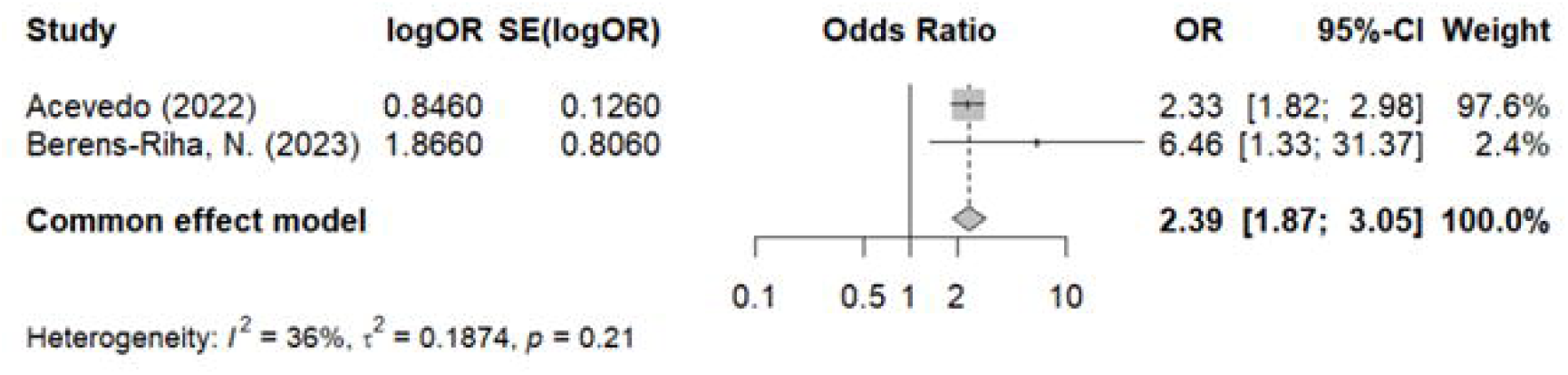

### Younger Age Group

Five studies (6, 41, 45, 49, 54), reported the younger age group as a risk factor for Mpox. We analyzed the data, and no evidence of heterogeneity was detected (*I*^2^ = 0, 0.47). Subsequently, a fixed-effects meta-analysis was employed and the results showed that the young age group was a risk factor for Mpox (OR = 2.03, 95%CI = 1.44, 2.85, P − value < 0.0001), **Figure 16**.

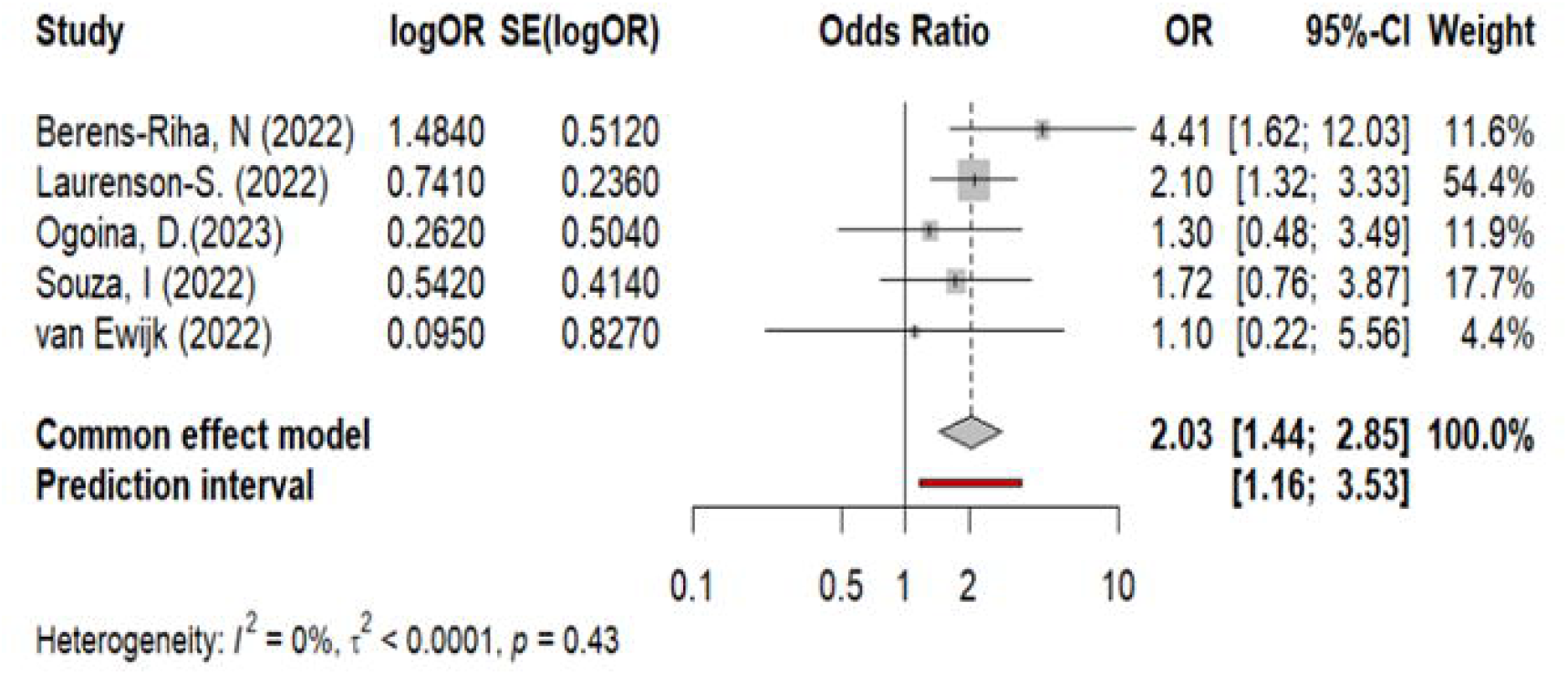

### Smallpox as a Protective Factor for Mpox

Two studies from Mpox endemic regions (6, 33) identified the smallpox vaccine as a protective factor against Mpox. The forest plot in **Figure 17** showed no evidence of heterogeneity (*I*^2^ = 0, 0.47), and subsequent meta-analysis using a fixed-effects model indicated a statistically significant negative association between Mpox and smallpox vaccination (OR = 0.24, 95%CI = 0.11, 0.55, P − value < 0.0001). This result suggested that the smallpox vaccination was a protective effect against Mpox infection.

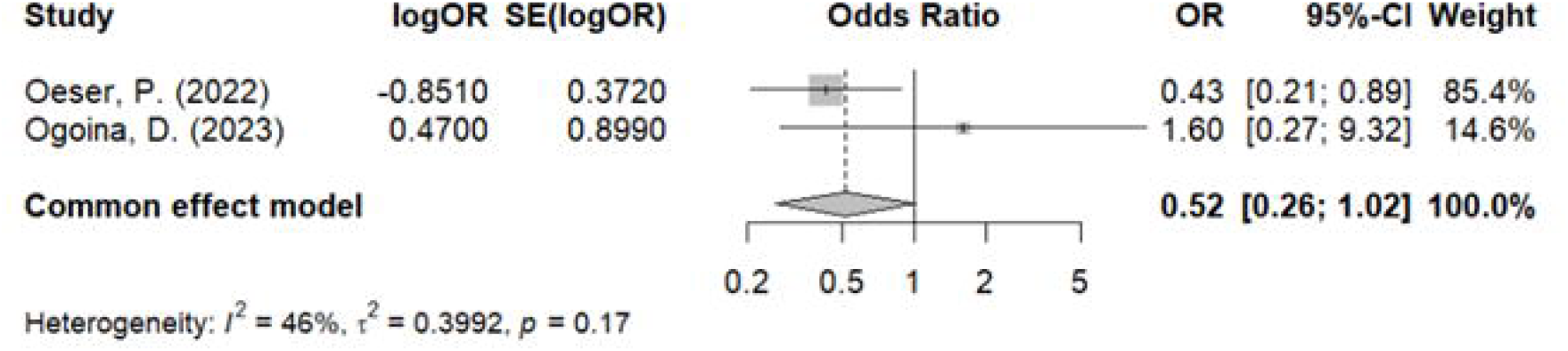

### Higher Educational Level as a Protective Factor for Mpox

Two studies (6, 55) reported that higher education level was a risk factor for Mpox. We analyzed the data, and moderate heterogeneity was detected (*I*^2^ = 46%, 0.17). Subsequently, a fixed-effects meta-analysis was employed, and according to the forest plot, a higher level of education had a negative influence on Mpox infection, however, no statistically significant result was found (OR = 0.52, 95%CI = 0.26, 1.02, P = 0.06), **Figure 18**.

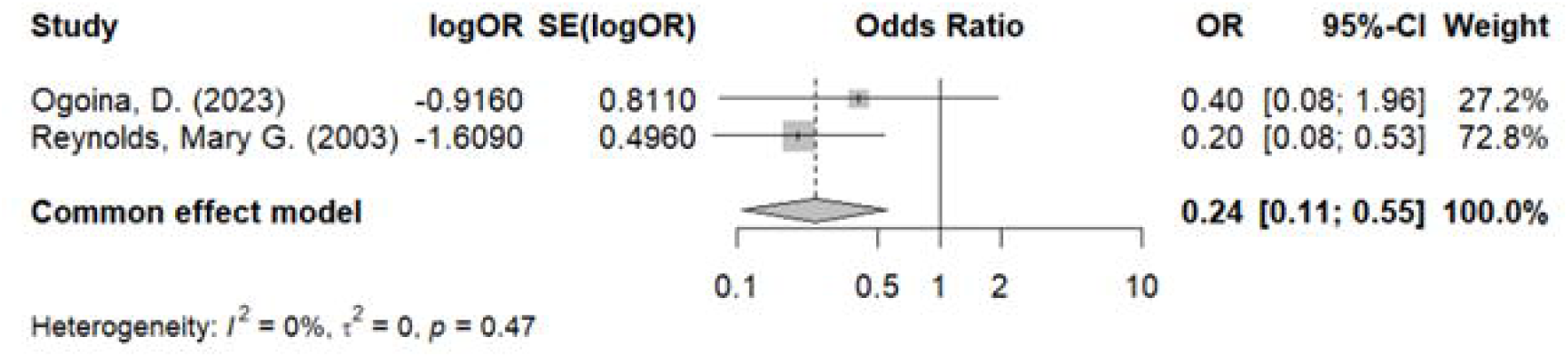

### Other Risk Factors

Other remaining risk factors identified in **Table 2** were only reported in one independent study, and therefore not analyzed. The details of these risk factors are shown in **Table 2**.

## Discussion

This systematic review and meta-analysis extracted data from published original articles of any study design (randomized control trial, case study, observational study, etc.) that investigated the risk factors of Mpox infection in human subjects and included 148,499 Mpox cases from 31 studies. These studies were rigorously chosen based on the NOS quality assessment to ensure the reliability of the evidence.

Findings from the current meta-analysis identified contact with infected animals, close contact with infected individuals, having multiple sexual partners, sexual contact, being identified as MSM (men who have sex with men), belonging to a younger age group, being HIV positive, having other sexually transmitted infections (STIs), and disease comorbidities as significant risk factors for Mpox infection. Conversely, smallpox vaccination emerged as a protective factor, particularly in endemic regions.

Contact with infected animals emerged as a significant risk factor for Mpox transmission, particularly in endemic regions such as the Democratic Republic of the Congo (DRC) and other parts of Central and Western Africa (26). The DRC reports most cases, where contact with animal reservoirs significantly drives Mpox infections (31). Anthropogenic and demographic changes since the 1980s may have increased local populations exposure to these reservoir species, elevating the risk of animal-to-human transmission (27, 30). In endemic regions, Mpox predominantly affects rural villages near tropical rainforests (29). According to Claire A. Quiner (29), humans primarily acquire Mpox through contact with infected animals or from limited human-to-human transmission chains. The virus has been isolated from wildlife only twice, highlighting the role of hunting and butchering bushmeat as primary zoonotic transmission activities (29, 30). These practices are widespread in Central Africa and are considered a primary route for several pathogens entering human populations. Findings by Mary G. Reynolds et al. (33) confirm that handling infected animals significantly correlates with Mpox infection, emphasizing the risk posed by direct contact with and exposure to excretions and secretions of infected animals. Adjusted data indicate a substantially higher likelihood of infection in individuals with daily exposure to, or direct contact with, sick animals (33, 57).

The meta-analysis revealed that HIV, other STIs, and the presence of comorbidities are significant risk factors for Mpox. HIV and other STIs are notably prevalent among these comorbidities, strongly influencing the risk of Mpox across various countries (6, 41, 45-47, 51, 55). This correlation highlights a crucial intersection of public health concerns, suggesting that efforts to control HIV and other STIs might also reduce Mpox susceptibility, especially in high-risk groups. These insights could inform broader healthcare strategies that concurrently address multiple infectious diseases, enhancing overall epidemic control.

Our study through meta-analysis identified having multiple sexual partners, close sexual contact, being identified as MSM, contact with a previously positive case, and belonging to a younger age group as significant risk factors for Mpox especially in non-endemic countries. Historically, Mpox cases have been linked to travel to Western and Central Africa where the disease is common, transmission from animals to humans via bodily fluids, and transmission between people via close contact with infectious sores or bodily fluids. (33, 34, 52). This is especially common among household members and healthcare professionals. However, recent findings suggest a shift towards person-to-person transmission, often through sexual contact, evidenced by the type and location of lesions that are primarily anal, rectal, or genital (58-60). This shift influenced a higher probability of Mpox transmission among younger individuals, particularly in non-endemic regions, who are identified as MSM group (61, 62).

Our meta-analysis found smallpox vaccination as a protective factor against Mpox. This protection may be attributed to immunity in older individuals who were previously vaccinated against smallpox (29, 33). Previous studies in endemic countries indicates that those vaccinated with the first-generation smallpox vaccine were less likely to develop severe Mpox (45, 63). Studies from the DRC also demonstrated the protect influence of smallpox vaccination among those born before the 1980s when smallpox was officially suspended (26, 31, 63).

### Strengths and limitations

The strength of our study lies in its robust methodology, which included a comprehensive literature search, study selection, clearly defined inclusion and exclusion criteria, eligibility screening, quality assessment, and the pooled analysis of Mpox data from 31 studies. Additionally, it is noteworthy that no significant unidentified heterogeneity persisted after synthesis. Our study has several limitations. First, no causal relationships between Mpox and risk factors were established. Second, the review included only English-language publications. Third, insufficient data prevented a meta-analysis of all risk factors, potentially due to fear of stigmatization in Mpox-endemic countries. However, the findings could aid clinicians and public health agencies in treatment decisions, supporting future research, and guiding policymakers in developing targeted interventions and control strategies.

## Conclusion

Our meta-analysis identified contact with infected animals, multiple sexual partners, sexual contact, younger age, HIV, other STIs, and comorbidities as significant risk factors for Mpox. We also found that the smallpox vaccine was protective against Mpox. As part of strategic control and prevention measures, it is important to prioritize laboratory testing for HIV and other STIs, especially since the recent outbreak among MSM was sustained by direct sexual contact. Additionally, extending Mpox vaccination to include high-risk groups engaging in sexual contact or intimate contact, even at mass gatherings, aligns with WHO guidelines and can reduce Mpox transmission globally.

## Supporting information

Quality Assessment Table

## Data Availability

All data produced in the present work are contained in the manuscript. Other information related to the present study are also available upon reasonable request to the authors.

## Ethics approval

This study was entirely based on published data. Therefore, an ethics committee approval or written informed consent was not required as no primary data was collected.

## Acknowledgments

W.A.W acknowledges financial support from the NSERC Discovery Grant (Appl No.: RGPIN-2023-05100).

## Supplementary Material

The original contributions presented in this study (NOS quality assessment) are included in the supplementary material of this article. For further inquiries, please contact the corresponding author.

## References

1. WHO. World Health Organization. Multi-country monkeypox outbreak: situation update [Internet].. Geneva: World Health Organization, 2022 [cited 2023 August 10] Available from https://wwwwhoint/emergencies/disease-outbreak-news/item/2022-DON390. 2022.

2. Kantele A, Chickering K, Vapalahti O, Rimoin A. Emerging diseases—the monkeypox epidemic in the Democratic Republic of the Congo. Clinical Microbiology and Infection. 2016;22(8):658–9.

3. Durski KN, McCollum AM, Nakazawa Y, Petersen BW, Reynolds MG, Briand S, et al. Emergence of monkeypox—west and central Africa, 1970--2017. Morbidity and mortality weekly report. 2018;67(10):306.

4. Eltvedt AK, Christiansen M, Poulsen A. A case report of monkeypox in a 4-year-old boy from the DR Congo: challenges of diagnosis and management. Case reports in pediatrics. 2020;2020.

5. Chieloka OS, Bammani IM, Amao LK. Descriptive epidemiology of the burden of human monkeypox in Nigeria: a retrospective review.

6. Ogoina D, Dalhat MM, Denue BA, Okowa M, Chika-Igwenyi NM, Yusuff HA, et al. Clinical characteristics and predictors of human mpox outcome during the 2022 outbreak in Nigeria: a cohort study. The Lancet Infectious Diseases. 2023;23(12):1418–28.

7. Moore MJ, Rathish B, Zahra F. Mpox (Monkeypox). Treasure Island (FL): StatPearls Publishing; 2022.

8. Bunge EM, Hoet B, Chen L, Lienert F, Weidenthaler H, Baer LR, et al. The changing epidemiology of human monkeypox—A potential threat? A systematic review. PLoS neglected tropical diseases. 2022;16(2):e0010141.

9. Dukers-Muijrers NHTM, Evers Y, Widdershoven V, Davidovich U, Adam PCG, Op de Coul ELM, et al. Mpox vaccination willingness, determinants, and communication needs in gay, bisexual, and other men who have sex with men, in the context of limited vaccine availability in the Netherlands (Dutch Mpox-survey). Frontiers in Public Health. 2023;10.

10. Kipkorir V, Dhali A, Srichawla B, Kutikuppala S, Cox M, Ochieng D, et al. The re-emerging monkeypox disease. Tropical Medicine & International Health. 2022;27(11):961--9.

11. Bragazzi NL, Woldegerima WA, Wu J, Converti M, Szarpak L, Crapanzano A, et al. Epidemiological and Clinical Characteristics of Mpox in Cisgender and Transgender Women and Non-Binary Individuals Assigned to the Female Sex at Birth: A Comprehensive, Critical Global Perspective. Viruses. 2024;16(2):325.

12. Delaney KP, Sanchez T, Hannah M, Edwards OW, Carpino T, Agnew-Brune C, et al. Strategies adopted by gay, bisexual, and other men who have sex with men to prevent monkeypox virus transmission—United States, August 2022. Morbidity and Mortality Weekly Report. 2022;71(35):1126.

13. Sukhdeo SS, Aldhaheri K, Lam PW, Walmsley S. A case of human monkeypox in Canada. CMAJ. 2022;194(29):E1031--E5.

14. Page MJ, McKenzie JE, Bossuyt PM, Boutron I, Hoffmann TC, Mulrow CD, et al. Updating guidance for reporting systematic reviews: development of the PRISMA 2020 statement. Journal of clinical epidemiology. 2021;134:103–12.

15. Shamseer L, Moher D, Clarke M, Ghersi D, Liberati A, Petticrew M, et al. Preferred reporting items for systematic review and meta-analysis protocols (PRISMA-P) 2015: elaboration and explanation. Bmj. 2015;349.

16. Hupe M. EndNote X9. Journal of Electronic Resources in Medical Libraries. 2019;16(3-4):117-9.

17. Deeks JJ, Higgins JP, Altman DG, Group CSM. Analysing data and undertaking meta-analyses. Cochrane handbook for systematic reviews of interventions. 2019:241–84.

18. Higgins JP, Thompson SG. Quantifying heterogeneity in a meta-analysis. Statistics in medicine. 2002;21(11):1539–58.

19. Hedges LV, Vevea JL. Fixed-and random-effects models in meta-analysis. Psychological methods. 1998;3(4):486.

20. Peters JL, Sutton AJ, Jones DR, Abrams KR, Rushton L. Comparison of two methods to detect publication bias in meta-analysis. Jama. 2006;295(6):676–80.

21. Islam RM, Oldroyd J, Karim MN, Hossain SM, Hoque DME, Romero L, et al. Systematic review and meta-analysis of prevalence of, and risk factors for, pelvic floor disorders in community-dwelling women in low and middle-income countries: a protocol study. BMJ open. 2017;7(6):e015626.

22. RSoftware. R: A language and environment for statistical computing. R Foundation for Statistical Computing, Vienna, Austria. 2021 [Available from: https://www.R-project.org/.

23. Balduzzi S, Rücker G, Schwarzer G. How to perform a meta-analysis with R: a practical tutorial. BMJ Ment Health. 2019;22(4):153–60.

24. Scale N-OQA. Case control studies. 2011.

25. Wells GA, Shea B, O’Connell D, Peterson J, Welch V, Losos M, et al. The Newcastle-Ottawa Scale (NOS) for assessing the quality of nonrandomised studies in meta-analyses. 2000.

26. Akilimali A, Adam MF, Awuah WA, Oduoye MO, Huang H. Human Monkey pox virus in Democratic Republic of the Congo: a potential health threat? IJS Global Health. 2023;6(2):e130.

27. Doshi RH, Guagliardo SAJ, Doty JB, Babeaux AD, Matheny A, Burgado J, et al. Epidemiologic and ecologic investigations of Monkeypox, Likouala department, Republic of the Congo, 2017. Emerging Infectious Diseases. 2019;25(2):273–81.

28. Nolen LD, Osadebe L, Katomba J, Likofata J, Mukadi D, Monroe B, et al. Introduction of monkeypox into a community and household: Risk factors and zoonotic reservoirs in the democratic republic of the congo. American Journal of Tropical Medicine and Hygiene. 2015;93(2):410–5.

29. Quiner CA, Moses C, Monroe BP, Nakazawa Y, Doty JB, Hughes CM, et al. Presumptive risk factors for monkeypox in rural communities in the Democratic Republic of the Congo. PloS one. 2017;12(2):e0168664.

30. Rimoin AW, Mulembakani PM, Johnston SC, Lloyd Smith JO, Kisalu NK, Kinkela TL, et al. Major increase in human monkeypox incidence 30 years after smallpox vaccination campaigns cease in the Democratic Republic of Congo. Proceedings of the National Academy of Sciences. 2010;107(37):16262–7.

31. Whitehouse ER, Bonwitt J, Hughes CM, Lushima RS, Likafi T, Nguete B, et al. Clinical and epidemiological findings from enhanced monkeypox surveillance in Tshuapa Province, Democratic Republic of the Congo during 2011–2015. The Journal of infectious diseases. 2021;223(11):1870–8.

32. Cline A, Marmon S. Demographics and disease associations of patients with mpox and recipients of mpox vaccine from safety net hospitals in New York City: A cross-sectional study. Journal of the American Academy of Dermatology. 2023;88(5):1160–3.

33. Reynolds MG, Davidson WB, Curns AT, Conover CS, Huhn G, Davis JP, et al. Spectrum of infection and risk factors for human monkeypox, United States, 2003. Emerging infectious diseases. 2007;13(9):1332.

34. Angelo KM, Smith T, Camprubí-Ferrer D, Balerdi-Sarasola L, Menéndez MD, Servera-Negre G, et al. Epidemiological and clinical characteristics of patients with monkeypox in the GeoSentinel Network: a cross-sectional study. The Lancet Infectious Diseases. 2023;23(2):196–206.

35. Vivancos R, Anderson C, Blomquist P, Balasegaram S, Bell A, Bishop L, et al. Community transmission of monkeypox in the United Kingdom, April to May 2022. Eurosurveillance. 2022;27(22):2200422.

36. Alegre B, Jubés S, Arango N, Pastene D, Lehrer E, Vilaseca I. Otorhinolaryngological manifestations in monkeypox. Acta Otorrinolaringologica Espanola. 2023;74(4):263–7.

37. Català A, Clavo-Escribano P, Riera-Monroig J, Martín-Ezquerra G, Fernandez-Gonzalez P, Revelles-Peñas L, et al. Monkeypox outbreak in Spain: clinical and epidemiological findings in a prospective cross-sectional study of 185 cases*. British Journal of Dermatology. 2022;187(5):765–72.

38. Estévez S, Vara M, Gamo M, Manzano S, Troya J, Botezat E, et al. Epidemiological and Clinical Characteristics of Patients Admitted to a Secondary Hospital with Suspected MPOX Virus Infection: Is HIV Playing a Role? Journal of Clinical Medicine. 2023;12(12).

39. García-Piqueras P, Bergón-Sendín M, Córdoba-García-Rayo M, Vírseda-González D, Medrano-Martínez N, Jiménez-Briones L, et al. Human monkeypox virus in a tertiary hospital in Madrid, Spain: an observational study of the clinical and epidemiological characteristics of 53 cases. Experimental Dermatology. 2023;32(2):198–202.

40. Martins-Filho PR, Nicolino RR, da Silva K. Incidence, geographic distribution, clinical characteristics, and socioeconomic and demographic determinants of monkeypox in Brazil: A nationwide population-based ecological study. Travel Medicine and Infectious Disease. 2023;52.

41. Souza INd, Pascom ARP, Spinelli MF, Dias GB, Barreira D, Miranda AE. Demographic and clinical characteristics of people diagnosed with active sexually transmitted infections among monkeypox cases in Brazil: the 2022 outbreak. Revista do Instituto de Medicina Tropical de São Paulo. 2024;66:e20.

42. Candela C, Raccagni AR, Bruzzesi E, Bertoni C, Rizzo A, Gagliardi G, et al. Human Monkeypox Experience in a Tertiary Level Hospital in Milan, Italy, between May and October 2022: Epidemiological Features and Clinical Characteristics. Viruses. 2023;15(3).

43. Ciccarese G, Di Biagio A, Bruzzone B, Guadagno A, Taramasso L, Oddenino G, et al. Monkeypox outbreak in Genoa, Italy: Clinical, laboratory, histopathologic features, management, and outcome of the infected patients. Journal of Medical Virology. 2023;95(2):e28560.

44. Dar NG, Alfaraj SH, Alboqmy KN, Amer H, Khanum N, Alshakrah F, et al. Locally acquired mpox outbreak in Riyadh, Saudi Arabia: clinical presentation, risk factors and preventive measures. Journal of Travel Medicine. 2023;30(8).

45. van Ewijk CE, Miura F, van Rijckevorsel G, de Vries HJ, Welkers MR, van den Berg OE, et al. Monkeypox outbreak in the Netherlands in 2022: public health response, epidemiological and clinical characteristics of the first 1000 cases and protection of the first-generation smallpox vaccine. medRxiv. 2022:2022.10. 20.22281284.

46. Zucker R, Lavie G, Wolff-Sagy Y, Gur-Arieh N, Markovits H, Abu-Ahmad W, et al. Risk assessment of human mpox infections: retrospective cohort study. Clinical Microbiology and Infection. 2023;29(8):1070–4.

47. Acevedo A, Garrido M. Epidemiological and clinical differences of confirmed and discarded Mpox cases on the 2022 Chilean outbreak. IJID Regions. 2023;9:59-62.

48. Alpalhão M, Sousa D, Frade JV, Patrocínio J, Garrido PM, Correia C, et al. Human immunodeficiency virus infection may be a contributing factor to monkeypox infection: analysis of a 42-case series. Journal of the American Academy of Dermatology. 2023;88(3):720–2.

49. Berens-Riha N, Bracke S, Rutgers J, Burm C, Van Gestel L, Hens M, et al. Persistent morbidity in Clade IIb mpox patients: interim results of a long-term follow-up study, Belgium, June to November 2022. Eurosurveillance. 2023;28(7).

50. Oeser P, Napierala H, Schuster A, Herrmann WJ. Risk Factors for Monkeypox Infection-a Cross-Sectional Study. Deutsches Arzteblatt international. 2023;120(5):65–6.

51. Lin M, Xin Y, Wang J, Nie P, Yan Q, Wang L, et al. Analysing monkeypox epidemic drivers: Policy simulation and multi-index modelling across 39 nations. Journal of Global Health. 2024;14.

52. Thornhill JP, Barkati S, Walmsley S, Rockstroh J, Antinori A, Harrison LB, et al. Monkeypox virus infection in humans across 16 countries—April–June 2022. New England Journal of Medicine. 2022;387(8):679–91.

53. Alhammadi OA, Al Hammadi A, Ganesan S, AlKaabi N, Al Harbi MS, Kamour AM, et al. Clinical characteristics of patients with mpox infection in the United Arab Emirates: a prospective cohort study. International Journal of Infectious Diseases. 2023;134:303–6.

54. Laurenson-Schafer H, Sklenovská N, Hoxha A, Kerr SM, Ndumbi P, Fitzner J, et al. Description of the first global outbreak of mpox: an analysis of global surveillance data. The Lancet Global Health. 2023;11(7):e1012–e23.

55. Oeser P, Napierala H, Schuster A, Herrmann WJ. Risk Factors for Monkeypox Infection—a Cross-Sectional Study. Deutsches Arzteblatt International. 2023;120(5):65–6.

56. Vallejo-Plaza A, Rodríguez-Cabrera F, Sebastián VH, Herrador BRG, Balader PS, Rodríguez-Alarcón LGSM, et al. Mpox (formerly monkeypox) in women: epidemiological features and clinical characteristics of mpox cases in Spain, April to November 2022. Eurosurveillance. 2022;27(48):2200867.

57. Doty JB, Malekani JM, Kalemba LN, Stanley WT, Monroe BP, Nakazawa YU, et al. Assessing monkeypox virus prevalence in small mammals at the human–animal interface in the democratic republic of the congo. Viruses. 2017;9(10).

58. Srivastava G, Srivastava G. Peculiar genital lesions. European Journal of Internal Medicine. 2023;113:100–1.

59. Derin O, Öztürk EN, Demirbaş ND, Sevgi DY, Dökmetaş İ. A Patient Presented with Genital Eruptions: The Second Case of Monkeypox from Türkiye. Mikrobiyoloji bulteni. 2023;57(1):134–40.

60. Pinnetti C, Mondi A, Mazzotta V, Vita S, Carletti F, Aguglia C, et al. Pharyngo-tonsillar involvement of Mpox in a cohort of men who have sex with men (MSM): A serious risk of missing diagnosis. Journal of Infection and Public Health. 2024;17(1):130–6.

61. Nimbi FM, Rosati F, Baiocco R, Giovanardi G, Lingiardi V. Monkeypox spread among men who have sex with men: how do people explain this relationship? A quali-quantitative study of beliefs among heterosexual and non-heterosexual Italian individuals. Psychology and Sexuality. 2023.

62. Movahedi Nia Z, Bragazzi N, Asgary A, Orbinski J, Wu J, Kong J. Mpox Panic, Infodemic, and Stigmatization of the Two-Spirit, Lesbian, Gay, Bisexual, Transgender, Queer or Questioning, Intersex, Asexual Community: Geospatial Analysis, Topic Modeling, and Sentiment Analysis of a Large, Multilingual Social Media Database. Journal of Medical Internet Research. 2023;25:e45108.

63. Rimoin AW, Mulembakani PM, Johnston SC, Lloyd Smith JO, Kisalu NK, Kinkela TL, et al. Major increase in human monkeypox incidence 30 years after smallpox vaccination campaigns cease in the Democratic Republic of Congo. Proceedings of the National Academy of Sciences. 2010;107(37):16262--7.

