## Supplementary material for "Risk Factors of Human Mpox (Monkeypox) Infection: A Systematic Review and Meta-Analysis": Quality Assessment Table

**Table 1: Studies Quality Assessment-New Castle Ottawa (NOS)**

| Study | Selection | Comparability | Exposure | Total |
| --- | --- | --- | --- | --- |
| Acevedo, A (2022) | ⭐⭐⭐ | ⭐⭐ | ⭐⭐⭐⭐ | 9 |
| Akilimali, A. (2022) | ⭐⭐⭐ | ⭐⭐ | ⭐⭐⭐ | 8 |
| Alegre, B. (2022) | ⭐⭐⭐ | ⭐⭐ | ⭐⭐ | 7 |
| Alhammadi, O. A. (2022) | ⭐⭐⭐ | ⭐ | ⭐⭐⭐ | 7 |
| Alpalhão, M. (2023) | ⭐⭐⭐ | ⭐⭐ | ⭐⭐ | 7 |
| Angelo, K. (2022) | ⭐⭐⭐⭐ | ⭐⭐ | ⭐⭐⭐ | 9 |
| Berens-Riha, N. (2022) | ⭐⭐⭐⭐ | ⭐⭐ | ⭐⭐⭐ | 9 |
| Candela, C (2022) | ⭐⭐⭐ | ⭐⭐ | ⭐⭐⭐ | 8 |
| Catala A (2022) | ⭐⭐⭐⭐ | ⭐⭐ | ⭐⭐⭐ | 9 |
| Ciccarese, G. (2022) | ⭐⭐⭐⭐ | ⭐⭐ | ⭐⭐⭐ | 9 |
| Cline, A. (2022) | ⭐⭐ | ⭐⭐ | ⭐⭐⭐ | 7 |
| Dar, N. G. (2023) | ⭐⭐ | ⭐⭐ | ⭐⭐⭐ | 7 |
| Doshi, R. H. (2017) | ⭐⭐ | ⭐⭐ | ⭐⭐⭐ | 7 |
| Estévez S. (2022) | ⭐⭐⭐⭐ | ⭐⭐ | ⭐⭐⭐ | 9 |
| García‐Piqueras (2022) | ⭐⭐⭐⭐ | ⭐⭐ | ⭐⭐⭐ | 9 |
| Laurenson-Schafer, (2023) | ⭐⭐⭐⭐ | ⭐⭐ | ⭐⭐⭐ | 9 |
| Lin, M. (2023) | ⭐⭐⭐⭐ | ⭐⭐ | ⭐⭐⭐ | 9 |
| Martins-Filho, P. R (2022) | ⭐⭐⭐ | ⭐ | ⭐⭐⭐ | 7 |
| Nolen, L. D (2015) | ⭐⭐⭐⭐ | ⭐⭐ | ⭐⭐⭐ | 9 |
| Oeser, P. (2022) | ⭐⭐⭐ | ⭐⭐ | ⭐⭐⭐⭐ | 9 |
| Ogoina, D. (2023) | ⭐⭐⭐ | ⭐⭐ | ⭐⭐⭐⭐ | 9 |
| Quiner, CA. (2016) | ⭐⭐⭐⭐ | ⭐⭐ | ⭐⭐ | 8 |
| Reynolds, Mary G (2003) | ⭐⭐⭐ | ⭐ | ⭐⭐⭐ | 7 |
| Rimoin, AW. (2010) | ⭐⭐⭐ | ⭐ | ⭐⭐⭐ | 7 |
| Souza I. (2022) | ⭐⭐⭐ | ⭐⭐ | ⭐⭐⭐ | 8 |
| Thornhill, J. (2022) | ⭐⭐⭐ | ⭐⭐ | ⭐⭐⭐ | 8 |
| Vallejo-Plaza (2022) | ⭐⭐⭐⭐ | ⭐⭐ | ⭐⭐ | 8 |
| van Ewijk (2022) | ⭐⭐⭐⭐ | ⭐ | ⭐⭐⭐ | 8 |
| Vivancos, Roberto | ⭐⭐⭐ | ⭐ | ⭐⭐⭐ | 7 |
| Whitehouse, E, | ⭐⭐⭐ | ⭐⭐ | ⭐⭐ | 7 |
| Zucker R. (2022) | ⭐⭐⭐ | ⭐⭐ | ⭐⭐⭐ | 8 |
